# Development and validation of PubMed and Ovid MEDLINE search filters for exposure pathways linking climate change with human health

**DOI:** 10.1101/2024.06.07.24308606

**Authors:** Maria-Inti Metzendorf, Ina Monsef, Katherine Jones, L Susan Wieland, Heidrun Janka, Camila Escobar Liquitay, Denise Thomson

## Abstract

**Introduction:** Climate change (CC) has major public and global health impacts to which policymakers need to respond. High-quality evidence syntheses (ES) are essential for policy-making. Search filters - validated combinations of search terms - play an important role in implementing robust search methods for ES. The identification of climate-health evidence presents challenges, such as the volume and multidisciplinary nature of the evidence and the fact that relevant studies do not consistently state their link to CC. Thus, our aim was to develop search filters for two search interfaces of the MEDLINE database.

**Methods:** CC impacts human health via several exposure pathways: extreme weather events, heat stress, air quality, water quality and quantity, food supply and safety, vector distribution and ecology, and social factors. We established a gold standard by comprehensively identifying health-related ES mentioning CC in five literature databases in February 2021. After screening 8,614 search results, we identified 110 ES for inclusion, extracted their included studies, and classified them according to exposure pathways. From this gold standard we empirically derived search terms per pathway and tested their performance with an independent set of studies.

**Results:** We extracted 2,324 studies from the first 79 ES. Based on a gold standard with 1,572 relevant studies indexed in PubMed, it was possible to develop and validate search filters with a sensitivity of 95%, 97% and 99% for six of the seven major climate-health exposure pathways. Filter development was not possible for one pathway due to the lack of coverage in MEDLINE.

**Conclusion:** We designed ready-to-use PubMed and Ovid MEDLINE search filters with a graded sensitivity for most exposure pathways linking CC with human health. These can be deployed by public and global health researchers conducting ES or primary research on climate-health to ensure robust identification of relevant evidence.

**KEY MESSAGES:**

- **What is already known on this topic**: The identification of evidence linking human health with climate change in literature databases such as MEDLINE presents challenges. Empirically derived search filters, which are validated combinations of search terms that can be readily used, are lacking for this topic.
- **What this study adds**: We present search filters with a graded sensitivity for six of the seven major climate-health exposure pathways (air quality, extreme weather events, food supply and safety, heat stress, vector distribution and ecology, water quality and quantity). A search filter for the pathway ‘social factors’ was not viable, suggesting that it requires other databases and complementary search methods to be used.
- **How this study might affect research, practice or policy**: The new search filters will help health researchers to identify relevant studies with a relationship to climate change. The filters can be applied independently from specific research questions (interventions, prognosis, associations, impacts, diseases, populations, or regions) as they focus on the major exposure pathways linking health with climate change.

## BACKGROUND

Climate change (CC) is considered the greatest public health challenge of the 21st century[1,2]. It causes long-term shifts in global or regional climate patterns, primarily characterized by alterations in temperature, precipitation, and other climatic factors attributed to the anthropogenic influence on greenhouse gas emissions (GHG), with profound impacts on ecosystems, human societies, and economies worldwide[1,3]. The climate crisis results in rising temperatures, rising sea levels and increased intensity, frequency, and duration of extreme weather events, which impact human health through a range of exposure pathways. Extreme weather events, heat stress, air quality, water quality and quantity, food supply and safety, vector distribution and ecology, and social factors are associated with multiple health outcomes such as injuries, heat stress, exacerbation of respiratory illnesses, increase of water-, food- and vector-borne diseases, adverse mental health impacts, forced migration and death[3,4].

Weather and climate variability have always played a role in affecting health impacts and outcomes[5]. The new challenge for policymakers is to iteratively manage adaptation to the constantly changing pressures of anthropogenic climate change. This will involve both managing the known impacts and, increasingly, incorporating the ability to prepare for unknown impacts to come[6]. Therefore, policymakers around the world must not only work within health systems on the local, regional and country-wide level, but also with other sectors of governance, such as municipalities, local communities and organizations, disaster response and natural resource authorities as well as agricultural departments to help protect populations from these impacts[7]. To do this effectively they must develop evidence-informed responses to both reduce the use of fossil fuels and GHG emissions and address the wide range of impacts that CC is having on all aspects of life on this planet, including human health and health systems[7,8].

High-quality evidence syntheses (ES) are a vital resource for policy-making, as a consolidated guide through a vast amount of pertinent policy-relevant research. They can range in scale from the massive reports produced by the Intergovernmental Panel on Climate Change (IPCC)[9] and the Lancet Countdown[1] to more focused publications such as systematic reviews or evidence gap maps[10]. ES can be used to collate information about the benefits, adverse effects and costs of interventions, as well as identifying gaps in knowledge and priorities for future research[11].

The quality of ES rests in part on the comprehensiveness of the included evidence, which in turn depends on the quality of the search strategies used to identify the evidence[12]. Developing search strategies for ES on the health impacts of CC can be complicated by the fact that relevant studies may not be labelled as being related to CC, the breadth of what can be considered as health impacts of CC, and the potential difficulty of teasing out health impacts distinct from social, economic and/or environmental impacts. In addition, the literature on the health-related aspects of the climate crisis covers several distinct fields: 1. impacts (effects on human health through several exposure pathways), 2. mitigation (interventions to reduce both the use of fossil fuels and greenhouse gas emissions, as well as the effects of these measures, including their health co-benefits), and 3. adaptation (interventions to adapt to the impacts of CC and reduce its detrimental effects on health, including vulnerability assessments and preventative approaches)[1,9].

The increasing number of ES being published on the health-related aspects of CC suggests a need for improved methods for their conduct[13]. Search filters – predefined and ideally empirically derived and validated combinations of search terms on a specific topic – play an important role in ensuring robust search methods for ES[14]. They are widely used by information specialists and medical librarians who support the search strategy design for ES. We searched the InterTASC Information Specialists’ sub-group search filters resource[15], the most comprehensive source for search filters, and did not find any published CC-specific search filters up to May 2024.

The aim of this methodological study is to describe the development and validation of search filters for established exposure pathways through which CC impacts human health. By focusing on the underlying pathways, we enable ES teams with different objectives to deploy them for a range of diverse synthesis topics, which can comprise the assessments of health impacts, the identification of preventative measures and vulnerable populations, supporting adaptation and resilience-building efforts, as well as policy prioritisation.

## METHODS

For developing search filters for exposure pathways through which human health is influenced by climate change, we focused on seven major pathways: air quality, extreme weather events, food supply and safety, heat stress, social factors, vector distribution and ecology, and water quality and quantity. The selection was based on established pathways linking climate change and health[3,8], while recognizing that the pathways and impacts might not be all-encompassing. Definitions and details of the exposure pathways are presented in Table 1.

**Table 1:**
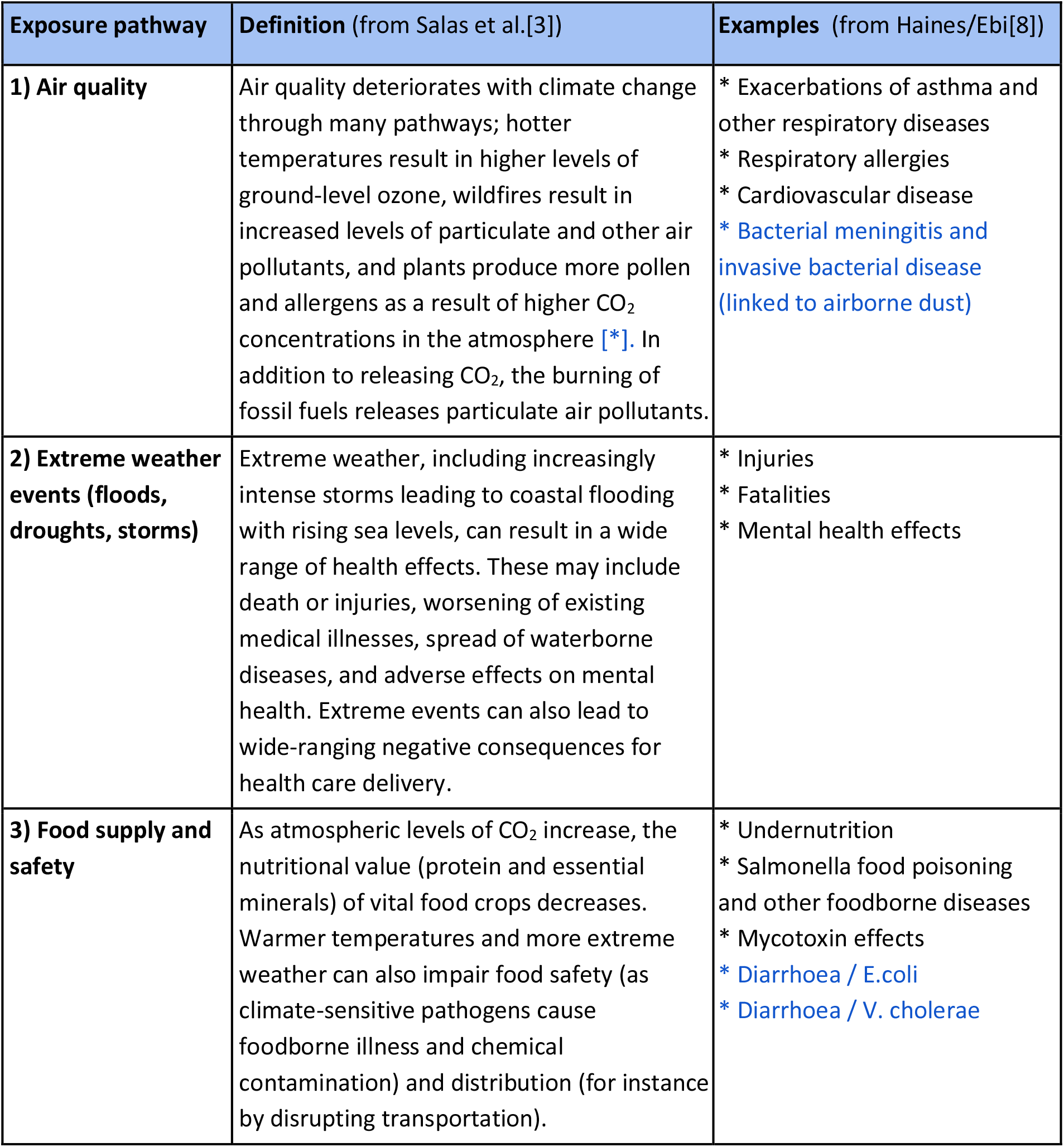
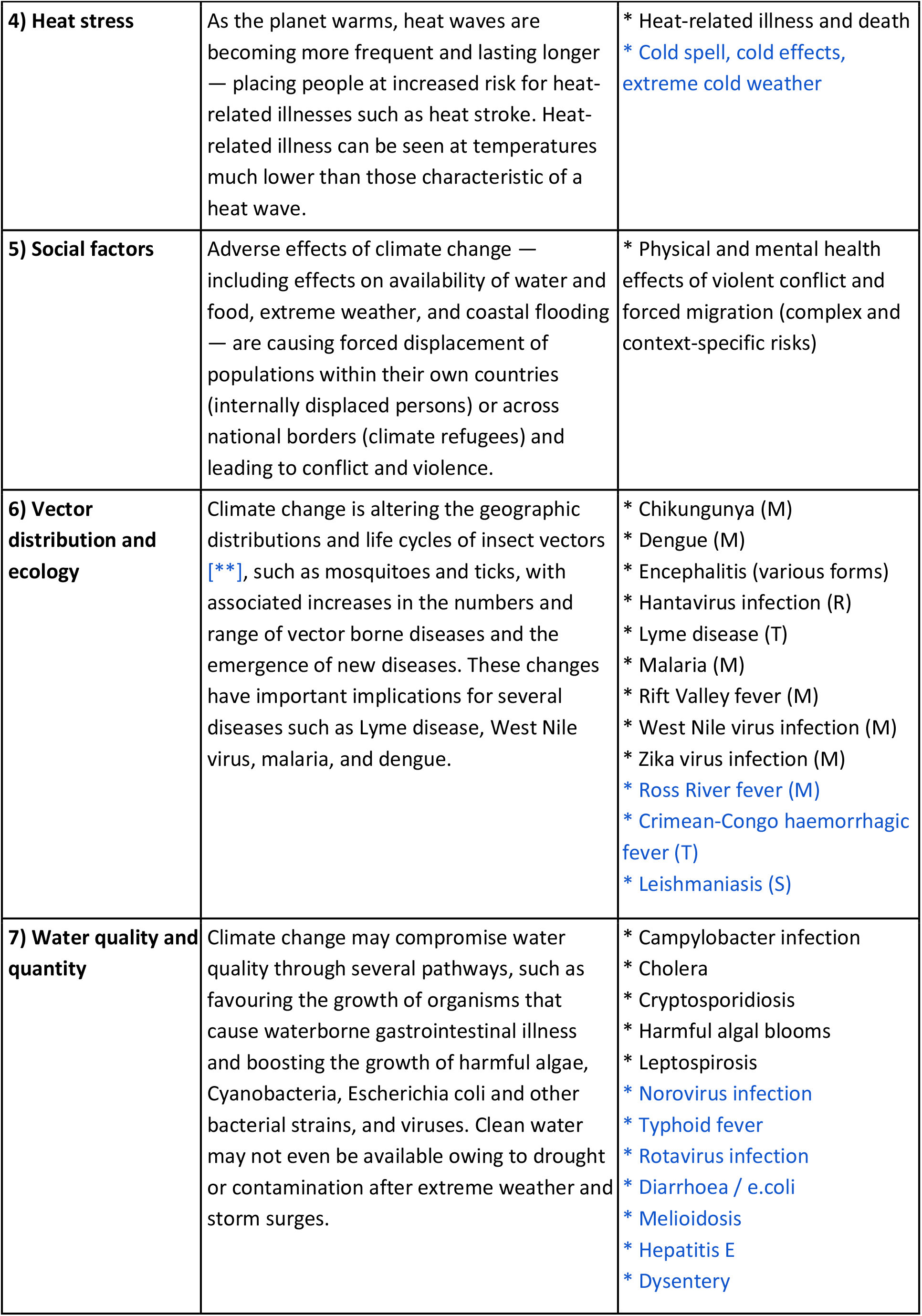
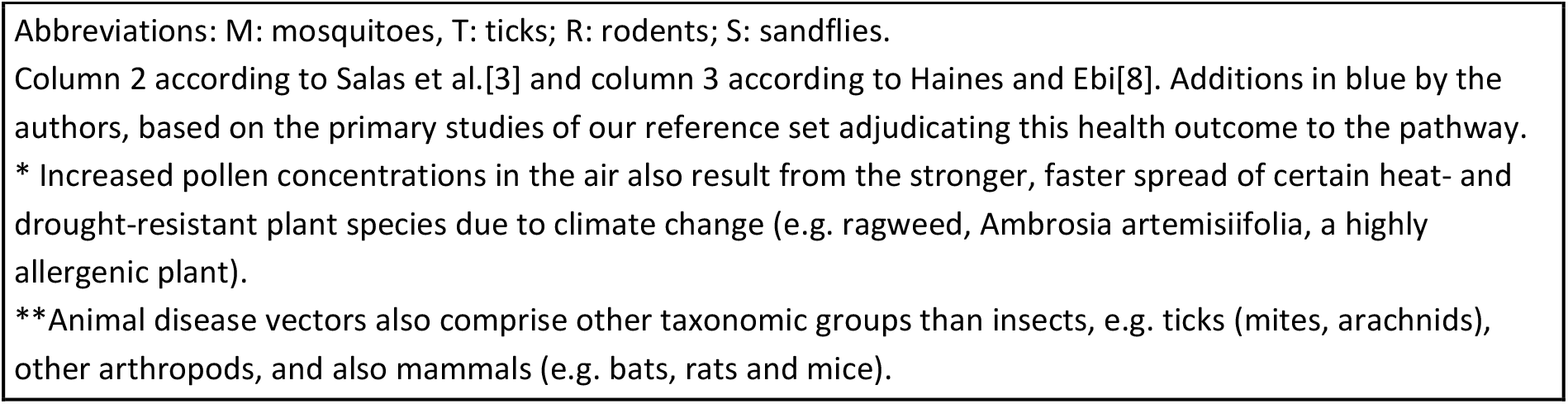
Exposure pathways through which human health is influenced by climate change.

| Exposure pathway | Definition (from Salas et al.[3]) | Examples (from Haines/Ebi[8]) |
| --- | --- | --- |
| **1) Air quality** | Air quality deteriorates with climate change through many pathways; hotter temperatures result in higher levels of ground-level ozone, wildfires result in increased levels of particulate and other air pollutants, and plants produce more pollen and allergens as a result of higher CO<sub>2</sub> concentrations in the atmosphere [*]. In addition to releasing CO<sub>2</sub>, the burning of fossil fuels releases particulate air pollutants. | * Exacerbations of asthma and other respiratory diseases<br>* Respiratory allergies<br>* Cardiovascular disease<br>* Bacterial meningitis and invasive bacterial disease (linked to airborne dust) |
| **2) Extreme weather events (floods, droughts, storms)** | Extreme weather, including increasingly intense storms leading to coastal flooding with rising sea levels, can result in a wide range of health effects. These may include death or injuries, worsening of existing medical illnesses, spread of waterborne diseases, and adverse effects on mental health. Extreme events can also lead to wide-ranging negative consequences for health care delivery. | * Injuries<br>* Fatalities<br>* Mental health effects |
| **3) Food supply and safety** | As atmospheric levels of CO<sub>2</sub> increase, the nutritional value (protein and essential minerals) of vital food crops decreases. Warmer temperatures and more extreme weather can also impair food safety (as climate-sensitive pathogens cause foodborne illness and chemical contamination) and distribution (for instance by disrupting transportation). | * Undernutrition<br>* Salmonella food poisoning and other foodborne diseases<br>* Mycotoxin effects<br>* Diarrhoea / E.coli<br>* Diarrhoea / V. cholerae |
| **4) Heat stress** | As the planet warms, heat waves are becoming more frequent and lasting longer — placing people at increased risk for heat-related illnesses such as heat stroke. Heat-related illness can be seen at temperatures much lower than those characteristic of a heat wave. | * Heat-related illness and death<br>* Cold spell, cold effects, extreme cold weather |
| **5) Social factors** | Adverse effects of climate change — including effects on availability of water and food, extreme weather, and coastal flooding — are causing forced displacement of populations within their own countries (internally displaced persons) or across national borders (climate refugees) and leading to conflict and violence. | * Physical and mental health effects of violent conflict and forced migration (complex and context-specific risks) |
| **6) Vector distribution and ecology** | Climate change is altering the geographic distributions and life cycles of insect vectors [**], such as mosquitoes and ticks, with associated increases in the numbers and range of vector borne diseases and the emergence of new diseases. These changes have important implications for several diseases such as Lyme disease, West Nile virus, malaria, and dengue. | * Chikungunya (M)<br>* Dengue (M)<br>* Encephalitis (various forms)<br>* Hantavirus infection (R)<br>* Lyme disease (T)<br>* Malaria (M)<br>* Rift Valley fever (M)<br>* West Nile virus infection (M)<br>* Zika virus infection (M)<br>* Ross River fever (M)<br>* Crimean-Congo haemorrhagic fever (T)<br>* Leishmaniasis (S) |
| **7) Water quality and quantity** | Climate change may compromise water quality through several pathways, such as favouring the growth of organisms that cause waterborne gastrointestinal illness and boosting the growth of harmful algae, Cyanobacteria, Escherichia coli and other bacterial strains, and viruses. Clean water may not even be available owing to drought or contamination after extreme weather and storm surges. | * Campylobacter infection<br>* Cholera<br>* Cryptosporidiosis<br>* Harmful algal blooms<br>* Leptospirosis<br>* Norovirus infection<br>* Typhoid fever<br>* Rotavirus infection<br>* Diarrhoea / e.coli<br>* Meloidosis<br>* Hepatitis E<br>* Dysentery |

Abbreviations: M: mosquitoes, T: ticks; R: rodents; S: sandflies.
Column 2 according to Salas et al.[3] and column 3 according to Haines and Ebi[8]. Additions in blue by the authors, based on the primary studies of our reference set adjudicating this health outcome to the pathway.
\* Increased pollen concentrations in the air also result from the stronger, faster spread of certain heat- and drought-resistant plant species due to climate change (e.g. ragweed, *Ambrosia artemisiifolia*, a highly allergenic plant).
\*\*Animal disease vectors also comprise other taxonomic groups than insects, e.g. ticks (mites, arachnids), other arthropods, and also mammals (e.g. bats, rats and mice).

### Building the gold standard (reference set)

To empirically derive search filters from the literature, we used the relative recall method[16] to establish a representative set of primary studies included in existing evidence syntheses (ES) on the health impacts of climate change (CC). Relative recall is based on the construction of a ‘gold standard’ (in the following termed ‘reference set’) of relevant studies on the search filter topic, which is divided into two groups. One group of study references, called the ‘development set’, is used to develop a search filter by analysing the text words (title and abstract) and the controlled vocabulary (,Medical Subject Headings’ assigned to references in the MEDLINE database), and the second set of references, called ‘validation set’, is used to test the performance of the developed search filter in retrieving those references[17].

Our methodological approach to constructing the reference, development, and validation sets for the search filters per exposure pathway is depicted in Figure 1 and described in the following paragraphs.

**Figure 1:**
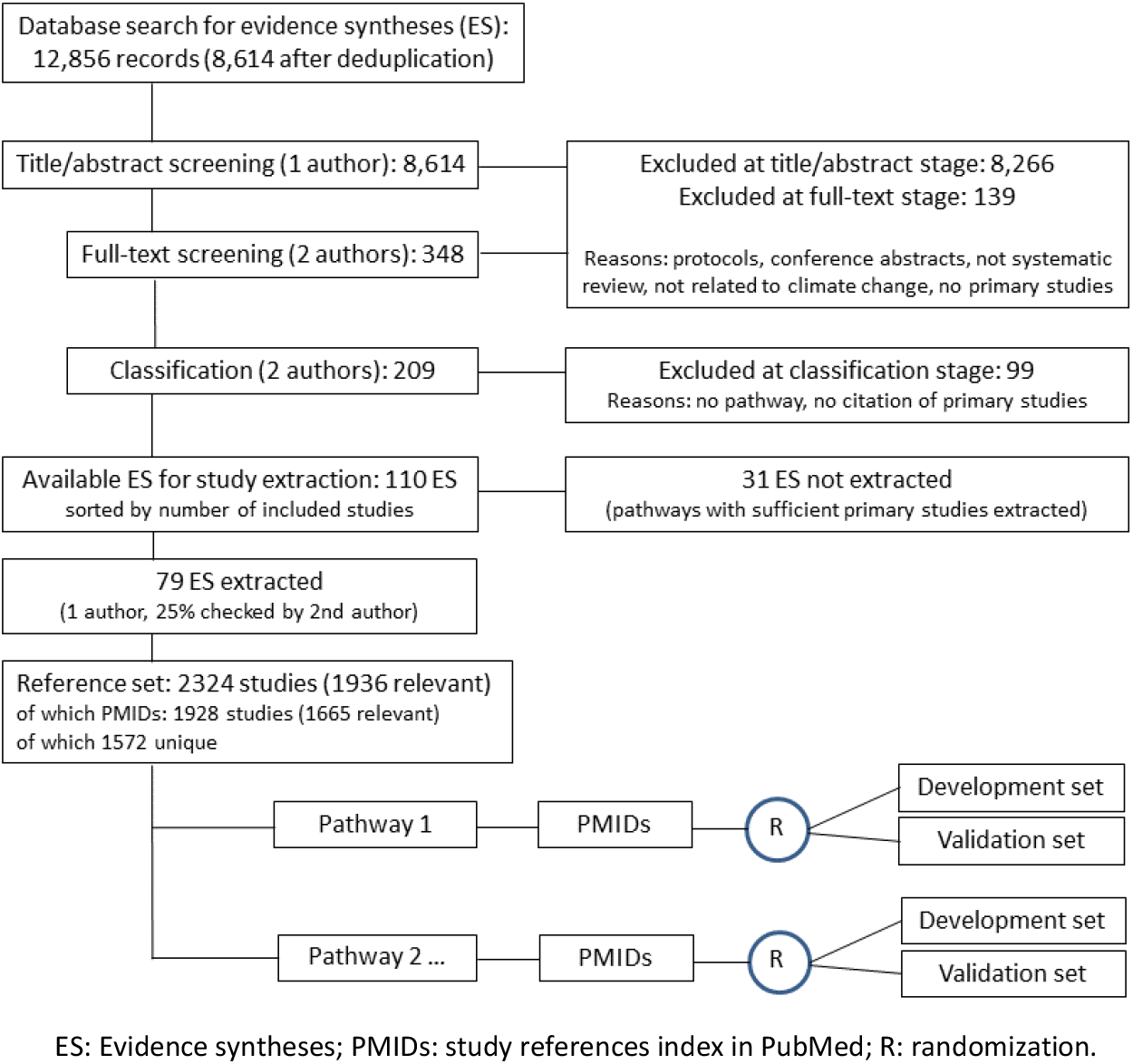
Methodological approach to construct the reference, development, and validation sets. ES: Evidence syntheses; PMIDs: study references index in PubMed; R: randomization.

#### Search for evidence syntheses

To identify potentially relevant ES, we used a sensitive search strategy developed by an experienced medical librarian and peer reviewed by an experienced health information specialist, both independent from the author team. This search strategy was run in MEDLINE, Embase, CINAHL, Cochrane Database of Systematic Reviews and ProQuest Dissertations and Theses on 10 February 2021 and was limited to English language ES published from 1999 onwards (for full details of the search strategy see Supplementary material - Appendix 1). The search yielded 12,856 records, which resulted in 8,614 records after deduplication in EndNote.

#### Inclusion and classification of evidence syntheses

The 8,614 references were screened on title/abstract-stage by one author (DT) using Covidence[18] according to the following inclusion criteria:

1. fully-published, health-related ES including primary studies (not a protocol or a conference abstract);
2. self-describes as ‘systematic (literature) review’, ‘scoping (literature) review’, ‘meta-analysis’, ‘comprehensive literature review’, ‘critical review’ in title or abstract, or is classified
3. as publication type ‘Review’ in the respective database and mentions ‘systematic (literature) search’ in the abstract;
4. mentions ‘climate change’, ‘global warming’, or ‘climate-health’ in title or abstract.

After the first round of screening, 348 references remained for inspection on the full-text stage. These records were re-assessed for the inclusion criteria by another author (KJ, MIM, IM, LSW) and discrepancies were resolved with a third author, who had not assessed the respective record. We excluded further 139 ES after assessment of the full-text.

The remaining 209 ES were divided in half and each half was extracted (bibliographic details, publication type) and classified per pathway by two independent authors (MIM+IM, KJ+LSW). Ten ES were piloted and discussed with the whole team before conducting the classification of the full set. If the ES covered more than one pathway, we classified it into all applicable pathways and if it did not fit into a pathway, we classified it as ‘miscellaneous’. This latter category was re-assessed and further classified by one author (DT) using free text words. In the last round, we further excluded 99 ES according to the following criteria:

1. thematic focus was not related to an exposure pathway (e.g. policies, research methods, professional roles, education, economics related to CC);
2. ES lacked sufficient information about the included studies (e.g. did not cite them).

This resulted in a final set of 110 relevant ES remaining for study extraction. Full details of the included ES are presented in Supplementary material - Appendix 2.

#### Extraction and classification of studies

Studies were extracted and classified by one member of the author team (with a few studies extracted by additional volunteers, see acknowledgements). However, during the classification process difficult cases were discussed in regular meetings of the author team and approximately 25% of the extracted studies were randomly rechecked by the first author (MIM).

We first extracted the number of included studies of each of the 110 identified relevant ES. To ensure maximum heterogeneity of topics, we then sorted them by their number of included studies (6 to 1,920) and started extracting the ES with the lowest number of studies. We extracted as many studies as possible for each pathway, but skipped study extraction of an ES after we had reached approximately 350 references for a pathway. We extracted each included study (bibliographic data, study design) and classified it into one or several exposure pathways. We also checked if the study was indexed in PubMed and recorded the PubMed ID (PMID) if available. If the study was not indexed in PubMed, we extracted the digital object identifier (DOI) if available. After finishing this process, we had extracted 79 ES published between 2008 and 2021, containing in total 2,324 studies. During classification, we categorized 388 of 2,324 studies as not relevant to our aim, for which the three main reasons were:

1. not a primary study (but an evidence synthesis);
2. not related to climate change (e.g. earthquakes, volcanic eruptions, air pollution caused by traffic, laboratory studies);
3. not adjudicable to a pathway.

This resulted in a set of 1,928 relevant references, of which 1,665 were indexed in PubMed. After removing duplicates, a final reference set of 1,572 unique references of primary studies indexed in PubMed was available for the generation of the development and validation sets. The resulting reference set is specified in Table 2.

**Table 2:** Reference set.

| Pathways | ES extracted (n) | References extracted (n) | PMIDs with duplicates (n) | Relevant PMIDs without duplicates (n) | References indexed in PubMed (%) |
| --- | --- | --- | --- | --- | --- |
| Air quality | 25 | 360 | 339 | 286 | 94.2 |
| Extreme weather events | 27 | 298 | 244 | 229 | 81.9 |
| Food supply and safety | 17 | 167 | 135 | 98 | 80.8 |
| Heat stress | 33 | 439 | 401 | 339 | 91.3 |
| Social factors | 7 | 73 | 29 | 28 | 39.7 |
| Vector distribution and ecology | 28 | 419 | 341 | 322 | 81.4 |
| Water quality and quantity | 20 | 392 | 351 | 270 | 89.5 |
| not relevant* | - | 388 | 263 | - | - |
| **Total\*\*** | **79** | **2,324\*\*\*** | **1,928** | **1,572** | **86.0** |
| - of those relevant | - | 1,936 | 1,665 | - | - |
| PMIDs: PubMed identifier.<br>*: 1. not a primary study; 2. not related to climate change; 3. not adjudicable to a pathway.<br>**: N of total ES/references (not sum of pathways, because classification into several pathways possible).<br>***: References with 2-5 pathways = 189 (175 with 2 pathways, 14 with >2 pathways). |  |  |  |  |  |

### Developing graded search filters per pathway

As proposed by Sampson and colleagues[16] a sample size of at least 100 references is required as a validation set for search filters, if the desired sensitivity is 0.9 (to establish a 95% confidence interval of .84 to .96). Therefore, we aimed to identify at least 200 references per pathway, to be able to split them in two sets and use 100 references for the validation.

After the reference set was established, we split the relevant references per pathway into two sets, a validation set (100 references) and a development set (the rest of available references per pathway). For this purpose, we put all unique identified references per pathway that were indexed in PubMed (PMIDs) in a spreadsheet column, randomised them using ‘random.org’[19], took the first 100 PMIDs as the validation set and used the rest of the references as the development set. This resulted in a development set of 186 PMIDs for ‘air quality’, 129 for ‘extreme weather events’, 239 for ‘heat stress’, 222 for ‘vector distribution and ecology’, and 170 for ‘water quality and quantity’. For two pathways we were not able to meet our objective of collecting at least 200 PMIDs. For ‘food supply and safety’ we only collected 98 PMIDs in total, and decided to use 60% as development set (58 PMIDs) and 40% as validation set (40 PMIDs). For the pathway ‘social factors’ we only collected 28 PMIDs in total. As we considered this to be insufficient, we decided against developing a search filter for this pathway.

We then proceeded to import the references of each pathway’s ‘development set’ into 1. Voyant Tools[20] for analysing phrases up to four words from the references’ titles and abstracts (tiab), and 2. into PubReMiner[21] for analysing single words from references’ titles and abstracts and the Medical Subject Headings (MeSH) associated with the references. We exported the outputs of each tool (frequencies of single words, phrases, and MeSH terms) into a Microsoft Excel spreadsheet for visual analysis and search term selection (separate columns for tiab-phrases, tiab-single words, and MeSH terms).

Two authors (MIM, IM) inspected: 1. all tiab-single terms that were included in at least five references of the development set and selected 25 potential terms for testing; 2. all tiab-phrases of two, three, and four words that were included in at least three references of the development set and selected 25 potential phrases for testing; 3. all MeSH-terms that were included in at least 20 references of the development set and selected 15 potential MeSH for testing with and without narrower terms. During selection, we chose the most frequent and at the same time most specific terms for each pathway.

We then proceeded to test all selected terms (single word, phrases, and MeSH) per pathway for their performance in retrieving the references of the respective development set (sensitivity) and recorded the number of hits they retrieved in PubMed. For each selected term, we then divided the number of hits retrieved in PubMed through its sensitivity. This gave us a proxy for the best performing terms, which we added incrementally to build a search string with the Boolean Operator ‘OR’ until we had reached a search string yielding the pre-defined cut-offs of 95%, 97% and 99% sensitivity in the development set. We then proceeded to validate the three graded search filters and calculated the sensitivity of all three search strings per pathway in the respective validation set.

#### Patient and public involvement

This study presents information retrieval methods for application by the public and global health research community as well as evidence synthesis teams, therefore patients or the public were not involved.

## RESULTS

Based on a dataset with 1,572 primary study references indexed in PubMed, it was possible to develop graded search filters with sensitivity cut-offs of 95%, 97% and 99% for six of the seven exposure pathways. The search filters and their performance indicators are presented in Table 3. For the exposure pathway ‘social factors’, we desisted from developing a search filter because we only identified 28 relevant references indexed in PubMed.

**Table 3:** Search filters per exposure pathway with their respective performance indicators.

|  | 95% sensitivity cut-off | 97% sensitivity cut-off | 99% sensitivity cut-off |
| --- | --- | --- | --- |
| **Air quality** |  |  |  |
| **S: development** | 96.2 | 97.8 | 99.5 |
| **S: validation** | 100 | 100 | 100 |
| **Hits** | 313,587 | 406,481 | 571,912 |
| **Filter** | WILDFIRE*[tiab] OR PM10[tiab] OR "pm 10"[tiab] OR Fires[mh] OR PM2[tiab] OR "air pollution"[tiab] OR "air quality"[tiab] OR "pm 2.5"[tiab] OR OZONE[tiab] OR "Particulate Matter"[mh] OR WEATHER[tiab] OR "Air Pollution"[mh] OR Smoke[mh] OR POLLUTANT*[tiab] OR HUMID*[tiab] | WILDFIRE*[tiab] OR PM10[tiab] OR "pm 10"[tiab] OR Fires[mh] OR PM2[tiab] OR "air pollution"[tiab] OR "air quality"[tiab] OR "pm 2.5"[tiab] OR OZONE[tiab] OR "Particulate Matter"[mh] OR WEATHER[tiab] OR "Air Pollution"[mh] OR Smoke[mh] OR POLLUTANT*[tiab] OR HUMID*[tiab] OR AMBIENT[tiab] | WILDFIRE*[tiab] OR PM10[tiab] OR "pm 10"[tiab] OR Fires[mh] OR PM2[tiab] OR "air pollution"[tiab] OR "air quality"[tiab] OR "pm 2.5"[tiab] OR OZONE[tiab] OR "Particulate Matter"[mh] OR WEATHER[tiab] OR "Air Pollution"[mh] OR Smoke[mh] OR POLLUTANT*[tiab] OR HUMID*[tiab] OR AMBIENT[tiab] OR ASTHMA[tiab] |
| **Extreme weather events** |  |  |  |
| **S: development** | 96.9 | 97.7 | 100 |
| **S: validation** | 91.0 | 95.0 | 97.0 |
| **Hits** | 279,064 | 292,897 | 435,499 |
| **Filter** | RIVER*[tiab] OR Floods[mh] OR "Cyclonic Storms"[mh] OR HURRICANE*[tiab] OR FLOOD*[tiab] OR DISASTER*[tiab] OR DROUGHT*[tiab] OR Disasters[mh] OR WEATHER[tiab] | RIVER*[tiab] OR Floods[mh] OR "Cyclonic Storms"[mh] OR HURRICANE*[tiab] OR FLOOD*[tiab] OR DISASTER*[tiab] OR DROUGHT*[tiab] OR Disasters[mh] OR WEATHER[tiab] OR RAINFALL*[tiab] | RIVER*[tiab] OR Floods[mh] OR "Cyclonic Storms"[mh] OR HURRICANE*[tiab] OR FLOOD*[tiab] OR DISASTER*[tiab] OR DROUGHT*[tiab] OR Disasters[mh] OR WEATHER[tiab] OR RAINFALL*[tiab] OR Climate[mh] |
| **Food supply and safety** |  |  |  |
| **S: development** | 96.6 | 98.3 | 100 |
| **S: validation** | 100 | 100 | 100 |
| **Hits** | 254,753 | 312,151 | 449,299 |
| **Filter** | "Child Nutrition Disorders"[mh] OR SALMONELLOS*[tiab] OR STUNT*[tiab] OR Cholera[mh] OR DROUGHT*[tiab] OR UNDERWEIGHT[tiab] OR Rain[mh] OR "Food Supply"[mh] OR FOODBORNE[tiab] OR Droughts[mh] OR MALNUTRITION[tiab] OR "Salmonella Infections"[mh] OR "Foodborne Diseases"[mh] OR WEATHER[tiab] | "Child Nutrition Disorders"[mh] OR SALMONELLOS*[tiab] OR STUNT*[tiab] OR Cholera[mh] OR DROUGHT*[tiab] OR UNDERWEIGHT[tiab] OR Rain[mh] OR "Food Supply"[mh] OR FOODBORNE[tiab] OR Droughts[mh] OR MALNUTRITION[tiab] OR "Salmonella Infections"[mh] OR "Foodborne Diseases"[mh] OR WEATHER[tiab] OR SALMONELLA*[tiab] | "Child Nutrition Disorders"[mh] OR SALMONELLOS*[tiab] OR STUNT*[tiab] OR Cholera[mh] OR DROUGHT*[tiab] OR UNDERWEIGHT[tiab] OR Rain[mh] OR "Food Supply"[mh] OR FOODBORNE[tiab] OR Droughts[mh] OR MALNUTRITION[tiab] OR "Salmonella Infections"[mh] OR "Foodborne Diseases"[mh] OR WEATHER[tiab] OR "Food Microbiology"[mh] OR SALMONELLA*[tiab] OR Seasons[mh] |
| **Heat stress** |  |  |  |
| **S: development** | 96.7 | 97.9 | 99.2 |
| **S: validation** | 98.0 | 98.0 | 98.0 |
| **Hits** | 445,025 | 588,595 | 715,676 |
| **Filter** | "Extreme Heat"[mh] OR "daily mortality"[tiab] OR "heat wave*"[tiab] OR "heat related"[tiab] OR "extreme heat"[tiab] OR HEATWAVE*[tiab] OR "Heat Stress Disorders"[mh] OR WEATHER[tiab] OR METEOROLOGIC*[tiab] OR "ambient temperature*"[tiab] OR "climate change"[tiab] OR "air pollution"[tiab] OR SUMMER*[tiab] OR "Hot Temperature"[mh] OR Seasons[mh] | "Extreme Heat"[mh] OR "daily mortality"[tiab] OR "heat wave*"[tiab] OR "heat related"[tiab] OR "extreme heat"[tiab] OR HEATWAVE*[tiab] OR "Heat Stress Disorders"[mh] OR WEATHER[tiab] OR METEOROLOGIC*[tiab] OR "ambient temperature*"[tiab] OR "climate change"[tiab] OR "air pollution"[tiab] OR SUMMER*[tiab] OR "Hot Temperature"[mh] OR CLIMATE*[tiab] OR Seasons[mh] OR HOT[tiab] | "Extreme Heat"[mh] OR "daily mortality"[tiab] OR "heat wave*"[tiab] OR "heat related"[tiab] OR "extreme heat"[tiab] OR HEATWAVE*[tiab] OR "Heat Stress Disorders"[mh] OR WEATHER[tiab] OR METEOROLOGIC*[tiab] OR "ambient temperature*"[tiab] OR "climate change"[tiab] OR "air pollution"[tiab] OR SUMMER*[tiab] OR "Hot Temperature"[mh] OR CLIMATE*[tiab] OR Seasons[mh] OR HOT[tiab] OR Climate[mh] OR "high temperature*"[tiab] OR AMBIENT[tiab] |
| **Vector distribution and ecology** |  |  |  |
| **S: development** | 96.8 | 97.3 | 99.1 |
| **S: validation** | 95.0 | 95.0 | 96.0 |
| **Hits** | 955,871 | 1,055,982 | 1,538,225 |
| **Filter** | RAIN*[tiab] OR Rain[mh:noexp] OR Climate[mh:noexp] OR "Insect Vectors"[mh:noexp] OR MOSQUITO*[tiab] OR "relative humidity"[tiab] OR "Climate Change"[mh:noexp] OR | RAIN*[tiab] OR Rain[mh:noexp] OR Climate[mh:noexp] OR "Insect Vectors"[mh:noexp] OR MOSQUITO*[tiab] OR "relative humidity"[tiab] OR "Climate Change"[mh:noexp] OR CLIMAT*[tiab] OR BORNE[tiab] OR | RAIN*[tiab] OR Rain[mh:noexp] OR Climate[mh:noexp] OR "Insect Vectors"[mh:noexp] OR MOSQUITO*[tiab] OR "Climate Change"[mh:noexp] OR CLIMAT*[tiab] OR HUMID*[tiab] OR BORNE[tiab] OR Seasons[mh:noexp] OR |
|  | CLIMAT*[tiab] OR<br>BORNE[tiab] OR<br>Seasons[mh:noexp] OR<br>ENDEMIC*[tiab] OR<br>transmission[sh:noexp]<br>OR "Disease<br>Outbreaks"[mh:noexp]<br>OR FEVER[tiab] | Seasons[mh:noexp] OR<br>ENDEMIC*[tiab] OR<br>transmission[sh:noexp] OR<br>"Disease<br>Outbreaks"[mh:noexp] OR<br>EPIDEMIC*[tiab] OR<br>FEVER[tiab] | ENDEMIC*[tiab] OR<br>transmission[sh:noexp] OR<br>"Disease<br>Outbreaks"[mh:noexp] OR<br>EPIDEMIC*[tiab] OR<br>FEVER[tiab] OR VECTOR*[tiab]<br>OR SURVEILLAN*[tiab] |
| **Water quality and quantity** |  |  |  |
| **S: development** | 95.9 | 97.1 | 99.4 |
| **S: validation** | 94.0 | 95.0 | 99.0 |
| **Hits** | 342,342 | 441,337 | 653,052 |
| **Filter** | "heavy rain*" [tiab] OR<br>Cholera[mh] OR<br>Floods[mh] OR "Vibrio<br>cholerae" [mh] OR<br>WATERBORNE[tiab] OR<br>RAINFALL* [tiab] OR<br>FLOOD* [tiab] OR<br>DIARRHEAL[tiab] OR<br>CHOLERA* [tiab] OR<br>"Water<br>Microbiology" [mh] OR<br>"Water Supply" [mh] OR<br>DIARRHOEA* [tiab] OR<br>Climate[mh] | "heavy rain*" [tiab] OR<br>Cholera[mh] OR Floods[mh]<br>OR "Vibrio cholerae" [mh] OR<br>WATERBORNE[tiab] OR<br>RAINFALL* [tiab] OR<br>FLOOD* [tiab] OR<br>DIARRH* [tiab] OR<br>CHOLERA* [tiab] OR "Water<br>Microbiology" [mh] OR<br>"Water Supply" [mh] OR<br>Diarrhea[mh] OR<br>Climate[mh] | "heavy rain*" [tiab] OR<br>Cholera[mh] OR Floods[mh]<br>OR "Vibrio cholerae" [mh] OR<br>WATERBORNE[tiab] OR<br>RAINFALL* [tiab] OR<br>FLOOD* [tiab] OR<br>DIARRH* [tiab] OR<br>CHOLERA* [tiab] OR "Water<br>Microbiology" [mh] OR "Water<br>Supply" [mh] OR Disasters[mh]<br>OR Climate[mh] OR<br>SEASON* [tiab] |
| S: sensitivity (defined as the number of relevant records in the development or validation set retrieved by the search filter divided by the total number of records in the development or validation set).<br>Hits: records retrieved by the respective filter in PubMed on 12.05.2024.<br>All search filters in PubMed syntax; Ovid MEDLINE syntax available as Supplementary Material - Appendix 3. |  |  |  |

The graded search filters were validated with an independent set of 100 references per pathway, except the search filters for the pathway ‘food supply and safety’, which were validated with 40 references. The validation showed that in some pathways the filters performed better (‘air quality’, ‘food supply and safety’, ‘heat stress’) than their indicated performance. In others (‘extreme weather events’, ‘vector distribution and ecology’, ‘water quality and quantity’), they performed slightly less well than their indicated performance, but the grading was reproducible.

With regard to the ‘hits in PubMed’ (records retrieved by the search filters on 12 May 2024), the filters yielded between 255,000 to 956,000 records in their 95% sensitivity cut-off versions and between 435,000 to 1,538,000 records in their 99% versions. This indicator can help researchers to choose a filter version in addition to considering its sensitivity. Of course, the number of records retrieved is dependent on the availability of literature in the respective pathway or ‘prevalence’ of the topic in PubMed. In practice, the search filters are intended to be combined with other concepts and/or their retrieved records will likely be further limited by geographic region, year range, or specific populations.

Interestingly, the indexing of the studies in PubMed showed a marked difference between the pathways, with three clusters emerging. Studies for the pathways ‘air quality’, ‘heat stress’, and ‘water quality and quantity’ had a very high indexing rate in PubMed between 90-94%, while studies for the pathways ‘extreme weather events’, ‘food supply and safety’, and ‘vector distribution and ecology’ had a less, but still good indexing rate in PubMed between 81-82%. The pathway ‘social factors’ had an exceptionally low indexing rate in PubMed of 40% (Table 2). This shows that some exposure pathways are well covered in PubMed, while others need to be supplemented by searching other databases and publication types, e.g. grey literature, or using complementary search methods, such as citation searching.

## DISCUSSION

Climate change (CC) poses significant threats to global public health, manifesting through various exposure pathways with complex and interconnected impacts. Recognizing the urgency of addressing these challenges, the United Nations Sustainable Development Goals (WHO/UN) underscore the imperative to mitigate CC and its adverse health effects[22]. As the body of evidence linking CC with health-related exposure pathways continues to grow, there is a pressing need for comprehensive evidence syntheses to inform policy-making at local, regional, and international levels. However, synthesising this expanding body of literature presents challenges, such as the volume of potentially relevant information, the fact that relevant primary studies do not consistently state the link to CC, and the multidisciplinary nature of the evidence.

Our aim was to develop robust and graded search filters for the major exposure pathways linking CC and health, which are essential for retrieving relevant literature. To our knowledge, this manuscript contributes the first search filters for climate-health exposure pathways. While it was possible to develop and validate search filters for five pathways as planned, for the pathway ‘food supply and safety’ we only had half the intended number of references available. Therefore, we are less confident that the graded sensitivities for this pathway are close to their true performance, which should be considered when using the search filter; we recommend using the 97% or 99% filter version rather than the 95% version. For the exposure pathway ‘social factors’, we suspended search filter development because we only identified 28 relevant references indexed in PubMed, suggesting that the identification of studies for this pathway requires other databases to be searched and complementary search methods to be used.

The main strengths of our search filters are that they will help researchers to identify relevant primary studies that do not mention their relationship to climate change, that they have been empirically and objectively developed by extracting and analysing the existing literature, and that they can be applied independently from specific research questions (interventions, prognosis, associations, impacts, diseases, populations, or regions) by focusing on the major exposure pathways linking health with CC.

Some limitations must be acknowledged. Firstly, the reliance on a single person for 3/4 of the data extraction and classification may have introduced bias or errors into the analysis. While difficult cases were discussed in the team, the possibility of misclassifications or overlooked pathways cannot be entirely discarded. Additionally, even though some of us have a public health background, we acknowledge our limited expertise in some of the pathway domains, which could also have resulted in erroneous or overlooked classifications. Last but not least, it would be beneficial to have an independent, external evaluation of our filters to help assess their performance in practice. We also expect that climate-health language will change. As terminology evolves and new concepts emerge, it will be essential to re-validate our search filters.

As Minx and colleagues argue, anthropogenic climate change is a wicked problem for which evidence-based policy responses are urgently needed[23]. Evidence syntheses presenting actionable messages for policymakers will support the learning processes that will enable initiatives to avoid, prepare for or manage the risks for health and other systems. Appropriate and relevant methods for such syntheses are needed, as the public and global health community needs to learn from other disciplines when tackling this work[23]. Thus, we recommend that author teams embarking on climate-health evidence syntheses consider the following: 1. use validated search filters to help capture relevant primary studies with high sensitivity and collaborate with medical librarians or other health information professionals when developing literature search strategies; 2. ensure they include people with expertise in climate change and, where needed, fields such as disaster research, equity, implementation science, etc.; 3. ensure they understand the differences between extreme weather events, climate variability and anthropogenic climate change; 4. prepare to incorporate sources of evidence not traditionally used in health-related evidence syntheses, such as non-randomised, economic and qualitative data, and even modelling studies in situations where direct evidence is not available[11]. Further research regarding information retrieval methods could include: 1. develop and validate climate-health search filters in different languages; 2. explore the possibility of constructing search filters for the pathways ‘food supply and safety’ and ‘social factors’ by examining the indexing of relevant publications in additional databases, such as the Web of Science Core Collection or the CABI Global Health database; 3. externally validate, review and update the search filters as the language on climate-health evolves and the evidence base expands.

## CONCLUSIONS

For six of seven main exposure pathways linking climate change and health, we developed and validated PubMed and Ovid MEDLINE search filters with a graded and high sensitivity, which can be readily used by public and global health researchers conducting evidence syntheses on climate-health. The availability of these filters represents a critical step towards helping to synthesize the growing body of evidence on the health impacts of climate change and enabling researchers to conduct syntheses with robust search methods that can explore the associations of climate change and health or inform policy-making and public health interventions addressing the mitigation of and adaptation to the developing climate emergency.

## Data Availability

All data relevant to the study are included in the article or the appendices. The full data sets of extracted studies and full details of the search filter development are available from the corresponding author upon request.

## CONTRIBUTIONS

Methods: MIM, DT, IM, KJ, LSW.

Screening and data extraction of evidence syntheses: IM, DT, MIM, KJ, LSW.

Data extraction and classification of primary studies: IM, KJ, MIM, DT, LSW, HJ, CEL.

Search filter development and validation: MIM, IM.

Drafting of manuscript: MIM.

Comments and review of manuscript draft: IM, KJ, DT, HJ, CEL, LSW.

## ACKNOWLEDGEMENTS

The search strategy was developed and run by Diana Keto-Lambert, MLIS, and peer-reviewed by Doug Salzwedel, MLIS. Data extraction of a small part of primary studies was aided by Radu-Florian Gherman and Raju Kanukula.

This work was first presented as an abstract in September 2023: Metzendorf MI, Monsef I, Jones K, Wieland LS, Escobar Liquitay C, Janka H, Thomson D: Understanding the health impacts of climate change: search filter development for exposure pathways. Abstracts accepted for the 27th Cochrane Colloquium, London, UK. Cochrane Database of Systematic Reviews 2023; (1 Supp 1): 37189.

## ETHICS APPROVAL

This study did not require ethics approval as it is a methodological study utilising published data. The data analysed were obtained from published evidence syntheses, ensuring compliance with ethical standards for research involving secondary data analysis. No new data were collected from human or animal subjects for this research.

