## Supplementary Material for "Development and validation of PubMed and Ovid MEDLINE search filters for exposure pathways linking climate change with human health"

**Appendix 1: Search strategies in electronic databases for identifying evidence syntheses related to climate change and health**

| **Date of search for all databases: 10 February 2021** | | | |
|---|---|---|---|
| **Database** | **Interface** | **Database Dates** | **Hits** |
| MEDLINE | Ovid | 1946 to Present | 4,413 |
| Embase | Ovid | 1974 to Present | 5,334 |
| CINAHL | Ebsco | 1970 to Present | 1,286 |
| Cochrane Database of Systematic Reviews | Wiley | Inception - Present | 200 |
| Dissertations & Theses | ProQuest | Inception - Present | 1,623 |
| Total search results: | | | **12,856** |
| Total search results after deduplication: | | | **8,614** |

| **Database: Ovid MEDLINE(R) and Epub Ahead of Print, In-Process & Other Non-Indexed Citations and Daily <1946 to February 09, 2021>** | | |
|---|---|---|
| 1 | exp climate change/ | 21323 |
| 2 | exp extreme weather/ | 169 |
| 3 | weather/ | 10567 |
| 4 | climate/ | 23356 |
| 5 | temperature/ | 244825 |
| 6 | climatic processes/ | 125 |
| 7 | cyclonic storms/ | 2319 |
| 8 | droughts/ | 8425 |
| 9 | floods/ | 2914 |
| 10 | greenhouse effect/ | 5738 |
| 11 | extreme heat/ | 410 |
| 12 | hot temperature/ | 117958 |

| | | |
|---|---|---|
| 13 | carbon footprint/ | 656 |
| 14 | ((climat* not (political-climate or organizational-climate)) adj3 (chang* or disruption* or instabilit* or stabilit* or stable or unstable or variable or variabilit* or vulnerab*)).tw,kf | 47817 |
| 15 | (weather adj3 (severe or chang* or disruption* or instabilit* or stabilit* or stable or unstable or variable or variabilit* or vulnerab*)).tw,kf. | 1822 |
| 16 | (environment* adj3 (chang* or disruption* or instabilit* or stabilit* or stable or unstable or variable or variabilit* or vulnerab*)).tw,kf. | 51412 |
| 17 | ((chang* or declin* or decreas* or increas*) adj3 humid*).tw,kf. | 2779 |
| 18 | ((global* or climate) adj2 (warm* or heat*)).tw,kf. | 12744 |
| 19 | climatic condition*.tw,kf. | 6453 |
| 20 | carbon footprint*.tw,kf. | 1057 |
| 21 | (greenhouse gas* or GHG or greenhouse effect* or carbon emission* or carbon dioxide emission* or CO2 emission*).tw,kf. | 15266 |
| 22 | (extreme-weather or extreme-heat or hot-weather or heat-wave* or heatwave or ((high* or hot* or warm*) adj3 temperature*)).tw,kf. | 97209 |
| 23 | (rain or rains or raining or precipitation or UV-index or flooding or floods or waterlogging or drought* or desertifcation or storm* or hailstorm* or hail or hailing* or monsoon* or typhoon*).tw,kf. | 131025 |
| 24 | (ice storm* or blizzard* or snowstorm* or extreme cold* or polar vortex*).tw,kf. | 1131 |
| 25 | (((high* or extreme) adj2 wind*) or hurricane* or cyclone* or tornado* or waterspout*).tw,kf. | 7658 |
| 26 | (forest fire* or wildfire* or wild fire*).tw,kf. | 3227 |
| 27 | (sea level* adj3 (rise or rises or rising or increase or increasing or increases)).tw,kf. | 1657 |
| 28 | or/1-27 [MeSH & KEYWORDS FOR CLIMATE CHANGE] | 668677 |
| 29 | Review Literature as Topic/ | 7860 |
| 30 | Review/ and scoping.tw,kf. | 5216 |

| | | |
|---|---|---|
| 31 | (scoping adj3 (review* or overview* or search* or study or studies or exercise* or project* or report* or methodolog*)).tw,kf. | 8916 |
| 32 | ((scope or scoping) adj2 evidence).tw,kf. | 136 |
| 33 | technical brief*.tw,kf. | 69 |
| 34 | ((evidence or systematic or literature or review* or overview*) adj3 map*).tw,kf. | 4195 |
| 35 | (structured adj2 literature review*).tw,kf. | 483 |
| 36 | ((Research or literature) adj3 (gap or gaps)).tw,kf. | 11408 |
| 37 | or/29-36 [TERMS FOR SCOPING REVIEWS] | 31282 |
| 38 | meta-analysis.pt. | 126311 |
| 39 | (meta-anal* or metaanal*).mp. | 227450 |
| 40 | ((quantitativ* adj3 review*) or (quantitativ* adj3 overview*)).mp. | 3988 |
| 41 | (quantitativ* adj3 synthes*).mp. | 3090 |
| 42 | (data synthes* or data extraction* or data abstraction*).mp. | 29886 |
| 43 | ((systematic* adj3 review*) or (systematic* adj3 overview*)).mp. | 233303 |
| 44 | ((methodologic adj3 review*) or (methodologic adj3 overview*)).mp. | 311 |
| 45 | (integrat* adj5 research).mp. | 10637 |
| 46 | (integrative adj3 (review* or overview*)).mp. | 4510 |
| 47 | (research adj3 (integrati* or overview*)).mp. | 5822 |
| 48 | (collaborative adj3 (review* or overview*)).mp. | 570 |
| 49 | (overview adj2 review*).mp. | 1275 |
| 50 | (rapid* adj2 review*).mp. | 1935 |
| 51 | or/38-50 [SYSTEMATIC, OVERVIEW OF, RAPID REVIEWS] | 381822 |
| 52 | review.pt. or (review* or overview*).mp. | 3894120 |
| 53 | (medline or medlars or pubmed or index medicus or embase or cochrane).mp. | 246441 |

| | | |
|---|---|---|
| 54 | (scisearch or web of science or psycinfo or psychinfo or cinahl or cinhal).mp. | 91794 |
| 55 | (excerpta medica or psychlit or psyclit or current contents or science citation index or sciences citation index or scopus).mp. | 34535 |
| 56 | (hand search* or handsearch* or manual search*).mp. | 14455 |
| 57 | ((electronic adj3 database*) or (bibliographic adj3 database*) or periodical index*).mp. | 42704 |
| 58 | (pooling or pooled or mantel haenszel).mp. | 115148 |
| 59 | (peto or der simonian or dersimonian or fixed effect*).mp. | 19320 |
| 60 | ((combine* or combining) adj5 (data or trial or trials or studies or study or result or results)).mp. | 131679 |
| 61 | or/53-60 | 500627 |
| 62 | 52 and 61 [MORE REVIEWS] | 270555 |
| 63 | Biomedical technology assessment/ | 10182 |
| 64 | (Technology adj1 (assessment* or overview* or appraisal*)).mp. | 15280 |
| 65 | (HTA or HTAs).mp. | 3494 |
| 66 | or/63-65 [HTAs] | 16722 |
| 67 | 37 or 51 or 62 or 66 [ALL REVIEWS] | 502595 |
| 68 | 28 and 67 [CLIMATE CHANGE AND REVIEWS] | 4729 |
| 69 | limit 68 to (english language and yr="1999 -Current") | 4502 |
| 70 | Case report/ | 2156467 |
| 71 | letter.pt. | 1123234 |
| 72 | historical article.pt. | 362127 |
| 73 | comment.pt. | 894000 |
| 74 | or/70-73 [FLUFF] | 3840897 |
| 75 | 69 not 74 | 4413 |

| Database: Embase <1974 to 2021 February 09> | | |
|---|---|---|
| 1 | exp climate change/ | 41143 |
| 2 | exp extreme weather/ | 312 |
| 3 | weather/ | 15013 |
| 4 | climate/ | 35833 |
| 5 | environmental temperature/ | 31275 |
| 6 | climatic processes/ | 31275 |
| 7 | drought/ | 11173 |
| 8 | Flooding/ | 7475 |
| 9 | greenhouse effect/ | 14510 |
| 10 | high temperature/ | 28880 |
| 11 | carbon footprint/ | 8145 |
| 12 | ((climat* not (political-climate or organizational-climate)) adj3 (chang* or disruption* or instabilit* or stabilit* or stable or unstable or variable or variabilit* or vulnerab*)).tw,kf. | 47808 |
| 13 | (weather adj3 (severe or chang* or disruption* or instabilit* or stabilit* or stable or unstable or variable or variabilit* or vulnerab*)).tw,kw. | 2124 |
| 14 | (environment* adj3 (chang* or disruption* or instabilit* or stabilit* or stable or unstable or variable or variabilit* or vulnerab*)).tw,kw. | 55067 |
| 15 | ((chang* or declin* or decreas* or increas*) adj3 humid*).tw,kw. | 3116 |
| 16 | ((global* or climate) adj2 (warm* or heat*)).tw,kw. | 1364 |
| 17 | climatic condition*.tw,kw. | 7053 |
| 18 | carbon footprint*.tw,kw. | 1377 |
| 19 | (greenhouse gas* or GHG or greenhouse effect* or carbon emission* or carbon dioxide emission* or CO2 emission*).tw,kw. | 1727 |
| 20 | (extreme-weather or extreme-heat or hot-weather or heat-wave* or heatwave or ((high* or hot* or warm*) adj3 temperature*)).tw,kw. | 93255 |

| | | |
|---|---|---|
| 21 | (rain or rains or raining or precipitation or UV-index or flooding or floods or waterlogging or drought* or desertifcation or storm* or hailstorm* or hail or hailing* or monsoon* or typhoon*).tw,kw. | 150989 |
| 22 | (ice storm* or blizzard* or snowstorm* or extreme cold* or polar vortex*).tw,kw. | 9199 |
| 23 | (((high* or extreme) adj2 wind*) or hurricane* or cyclone* or tornado* or waterspout*).tw,kw. | 9199 |
| 24 | (forest fire* or wildfire* or wild fire*).tw,kw. | 3603 |
| 25 | (sea level* adj3 (rise or rises or rising or increase or increasing or increases)).tw,kw. | 1574 |
| 26 | or/1-25 [MeSH & KEYWORDS FOR CLIMATE CHANGE] | 437680 |
| 27 | "review"/ and scoping.tw,kw. | 3283 |
| 28 | (scoping adj3 (review* or overview* or search* or study or studies or exercise* or project* or report* or methodolog*)).tw,kw. | 9472 |
| 29 | ((scope or scoping) adj2 evidence).tw,kw. | 159 |
| 30 | technical brief*.tw,kw. | 63 |
| 31 | ((evidence or systematic or literature or review* or overview*) adj3 map*).tw,kw. | 4843 |
| 32 | (structured adj2 literature review*).tw,kw. | 656 |
| 33 | ((Research or literature) adj3 (gap or gaps)).tw,kw. | 12860 |
| 34 | or/27-33 [TERMS FOR SCOPING REVIEWS] | 26544 |
| 35 | meta-analysis.pt. | 0 |
| 36 | (meta-anal* or metaanal*).mp. | 329359 |
| 37 | ((quantitativ* adj3 review*) or (quantitativ* adj3 overview*)).mp. | 4651 |
| 38 | (quantitativ* adj3 synthes*).mp. | 3556 |
| 39 | (data synthes* or data extraction* or data abstraction*).mp. | 45896 |
| 40 | ((systematic* adj3 review*) or (systematic* adj3 overview*)).mp. | 381875 |

| | | |
|---|---|---|
| 41 | ((methodologic adj3 review*) or (methodologic adj3 overview*)).mp. | 356 |
| 42 | (integrat* adj5 research).mp. | 13449 |
| 43 | (integrative adj3 (review* or overview*)).mp. | 4642 |
| 44 | (research adj3 (integrati* or overview*)).mp. | 7023 |
| 45 | (collaborative adj3 (review* or overview*)).mp. | 792 |
| 46 | (overview adj2 review*).mp. | 1386 |
| 47 | (rapid* adj2 review*).mp. | 2264 |
| 48 | or/35-47 [SYSTEMATIC, OVERVIEW OF, RAPID REVIEWS] | 578463 |
| 49 | review.pt. or (review* or overview*).mp. | 4726833 |
| 50 | (medline or medlars or pubmed or index medicus or embase or cochrane).mp. | 323362 |
| 51 | (scisearch or web of science or psycinfo or psychinfo or cinahl or cinhal).mp. | 103149 |
| 52 | (excerpta medica or psychlit or psyclit or current contents or science citation index or sciences citation index or scopus).mp. | 49866 |
| 53 | (hand search* or handsearch* or manual search*).mp. | 17480 |
| 54 | ((electronic adj3 database*) or (bibliographic adj3 database*) or periodical index*).mp. | 57246 |
| 55 | (pooling or pooled or mantel haenszel).mp. | 161712 |
| 56 | (peto or der simonian or dersimonian or fixed effect*).mp. | 25722 |
| 57 | ((combine* or combining) adj5 (data or trial or trials or studies or study or result or results)).mp. | 165888 |
| 58 | or/50-57 | 658794 |
| 59 | 49 and 58 [MORE REVIEWS] | 355960 |
| 60 | Biomedical technology assessment/ | 14946 |
| 61 | (Technology adj1 (assessment* or overview* or appraisal*)).mp. | 22382 |
| 62 | (HTA or HTAs).mp. | 7747 |

| | | |
|---|---|---|
| 63 | or/60-62 [HTAs] | 26353 |
| 64 | Case report/ | 2585669 |
| 65 | Letter/ | 1098841 |
| 66 | Note/ | 789512 |
| 67 | Editorial/ | 666099 |
| 68 | or/64-67 [FLUFF] | 4857734 |
| 69 | 34 or 48 or 59 or 63 [ALL REVIEW TYPES] | 717061 |
| 70 | 26 and 69 [CLIMATE CHANGE AND ALL REVIEWS] | 5694 |
| 71 | 70 not 68 [FLUFF REMOVED] | 5569 |
| 72 | limit 71 to (english language and yr="1999 -Current") | 5334 |

| **Database: CINAHL Plus Full Text** | | |
|---|---|---|
| S61 | S22 AND S60 Limiters - English Language; Published Date: 19990101-20210231 | 1,276 |
| S60 | S31 OR S45 OR S56 OR S59 | 239,018 |
| S59 | S57 OR S58 | 818 |
| S58 | (HTA or HTAs) | 1,349 |
| S57 | (Technology n1 (assessment* or overview* or appraisal*)) | 3,875 |
| S56 | S46 AND S55 | 133,869 |
| S55 | S47 OR S48 OR S49 OR S50 OR S51 OR S52 OR S53 OR S54 | 208,737 |
| S54 | ((combine* or combining) n5 (data or trial or trials or studies or study or result or results)) | 23,735 |
| S53 | (peto or "der simonian" or dersimonian or "fixed effect*") | 6,718 |
| S52 | (pooling or pooled or "mantel haenszel") | 34,580 |

| | | |
|---|---|---|
| S51 | ((electronic n3 database*) or (bibliographic n3 database*) or periodical index*) | 18,456 |
| S50 | ("hand search*" or handsearch* or "manual search*") | 6,244 |
| S49 | (excerpta medica or psychlit or psyclit or current contents or science citation index or sciences citation index or scopus) | 26,378 |
| S48 | (scisearch or web of science or psycinfo or psychinfo or cinahl or cinhal) | 73,137 |
| S47 | (medline or medlars or pubmed or index medicus or embase or cochrane) | 132,350 |
| S46 | (review* or overview*) | 751,091 |
| S45 | S32 OR S33 OR S34 OR S35 OR S36 OR S37 OR S38 OR S39 OR S40 OR S41 OR S42 OR S43 OR S44 | 197,434 |
| S44 | (rapid* n2 review*) | 975 |
| S43 | (overview n2 review*) | 2,127 |
| S42 | (collaborative n3 (review* or overview*)) | 393 |
| S41 | (research n3 (integrati* or overview*)) | 3,933 |
| S40 | (integrative n3 (review* or overview*)) | 5,548 |
| S39 | (integrat* n5 research) | 5,714 |
| S38 | ((methodologic n3 review*) or (methodologic n3 overview*)) | 168 |
| S37 | ((systematic* n3 review*) or (systematic* n3 overview*)) | 148,654 |
| S36 | ("data synthes*" or "data extraction*" or "data abstraction*") | 11,249 |
| S35 | (quantitativ* n3 synthes*) | 1,054 |
| S34 | ((quantitativ* n3 review*) or (quantitativ* n3 overview*)) | 2,174 |
| S33 | (meta-anal* or metaanal*) | 89,681 |
| S32 | (MH "Meta Analysis") | 51,898 |
| S31 | S23 OR S24 OR S25 OR S26 OR S27 OR S28 OR S29 OR S30 | 21,803 |
| S30 | TI ( ((Research or literature) n3 (gap or gaps)) ) OR AB ( ((Research or literature) n3 (gap or gaps)) ) | 6,884 |

| | | |
|---|---|---|
| S29 | TI (structured n2 "literature review*") OR AB (structured n2 "literature review*") | 301 |
| S28 | TI ( ((evidence or systematic or literature or review* or overview*) n3 map*) ) OR AB ( ((evidence or systematic or literature or review* or overview*) n3 map*) ) | 1,550 |
| S27 | TI "technical brief*" OR AB "technical brief*" | 55 |
| S26 | TI ( ((scope or scoping) n2 evidence) ) OR AB ( ((scope or scoping) n2 evidence) ) | 178 |
| S25 | TI ( (scoping n3 (review* or overview* or search* or study or studies or exercise* or project* or report* or methodolog*)) ) OR AB ( (scoping n3 (review* or overview* or search* or study or studies or exercise* or project* or report* or methodolog*)) ) | 5,260 |
| S24 | (MH "Scoping Review") | 2,589 |
| S23 | (MH "Literature Review") | 8,317 |
| S22 | S1 OR S2 OR S3 OR S4 OR S5 OR S6 OR S7 OR S8 OR S9 OR S10 OR S11 OR S12 OR S13 OR S14 OR S15 OR S16 OR S17 OR S18 OR S19 OR S20 OR S21 | 50,034 |
| S21 | TI ( ("sea level*" n3 (rise or rises or rising or increase or increasing or increases)) ) OR AB ( ("sea level*" n3 (rise or rises or rising or increase or increasing or increases)) ) | 168 |
| S20 | TI ( ("forest fire*" or wildfire* or "wild fire*") ) OR AB ( ("forest fire*" or wildfire* or "wild fire*") ) | 593 |
| S19 | TI ( (((high* or extreme) adj2 wind*) or hurricane* or cyclone* or tornado* or waterspout*) ) OR AB ( (((high* or extreme) adj2 wind*) or hurricane* or cyclone* or tornado* or waterspout*)) | 3,291 |
| S18 | TI ( ("ice storm*" or blizzard* or snowstorm* or "extreme cold*" or "polar vortex*") ) OR AB ( ("ice storm*" or blizzard* or snowstorm* or "extreme cold*" or "polar vortex*") ) | 245 |
| S17 | TI ( (rain or rains or raining or precipitation or UV-index or flooding or floods or waterlogging or drought* or desertifcation or storm* or hailstorm* or hail or hailing* or monsoon* or typhoon*) ) OR AB ( (rain or rains or raining or precipitation or UV-index or flooding or floods or waterlogging or drought* or desertifcation or storm* or hailstorm* or hail or hailing* or monsoon* or typhoon*) ) | 8,673 |

| | | |
|---|---|---|
| S16 | TI ( ("extreme weather" or "extreme heat" or "hot weather" or "heat wave*" or heatwave or ((high* or hot* or warm*) n3 temperature*)) ) OR AB ( ("extreme weather" or "extreme heat" or "hot weather" or "heat wave*" or heatwave or ((high* or hot* or warm*) n3 temperature*)) ) | 4,384 |
| S15 | TI ( ("greenhouse gas*" or GHG or "greenhouse effect*" or "carbon emission*" or "carbon dioxide emission*" or "CO2 emission*") ) OR AB ( ("greenhouse gas*" or GHG or "greenhouse effect*" or "carbon emission*" or "carbon dioxide emission*" or "CO2 emission*") ) | 948 |
| S14 | TI "carbon footprint*" OR AB "carbon footprint*" | 244 |
| S13 | TI "climatic condition*" OR AB "climatic condition*" | 221 |
| S12 | TI ( ((global* or climate) n2 (warm* or heat*)) ) OR AB ( ((global* or climate) n2 (warm* or heat*)) ) | 1,029 |
| S11 | TI ( ((chang* or declin* or decreas* or increas*) n3 humid*) ) OR AB ( ((chang* or declin* or decreas* or increas*) n3 humid*) ) | 248 |
| S10 | TI ( (environment* n3 (chang* or disruption* or instabilit* or stabilit* or stable or unstable or variable or variabilit* or vulnerab*)) ) OR AB ( (environment* n3 (chang* or disruption* or instabilit* or stabilit* or stable or unstable or variable or variabilit* or vulnerab*)) ) | 8,977 |
| S9 | TI ( (weather n3 (severe or chang* or disruption* or instabilit* or stabilit* or stable or unstable or variable or variabilit* or vulnerab*)) ) OR AB ( (weather n3 (severe or chang* or disruption* or instabilit* or stabilit* or stable or unstable or variable or variabilit* or vulnerab*)) | 455 |
| S8 | TI ( ((climat* not ("political climate" or "organizational climate")) n3 (chang* or disruption* or instabilit* or stabilit* or stable or unstable or variable or variabilit* or vulnerab*)) ) OR AB ( ((climat* not ("political climate" or "organizational climate")) n3 (chang* or disruption* or instabilit* or stabilit* or stable or unstable or variable or variabilit* or vulnerab*)) ) | 5,093 |
| S7 | (MH "Carbon Footprint") | 81 |
| S6 | (MH "Greenhouse Effect") | 1,196 |
| S5 | (MH "Natural Disasters") | 12,009 |
| S4 | (MH "Temperature") | 7,679 |
| S3 | (MH "Climate") | 3,403 |

| | | |
|---|---|---|
| S2 | (MH "Weather") | 3,017 |
| S1 | (MH "Climate Change") | 1,936 |

| **Database: The Cochrane Library** | | |
|---|---|---|
| #1 | [mh "climate change"] | 7 |
| #2 | [mh "extreme weather"] | 1 |
| #3 | [mh ^weather] | 39 |
| #4 | [mh ^climate] | 54 |
| #5 | [mh ^temperature] | 1352 |
| #6 | [mh ^"climatic processes"] | 0 |
| #7 | [mh ^"cyclonic storms"] | 5 |
| #8 | [mh ^"droughts"] | 2 |
| #9 | [mh ^floods] | 4 |
| #10 | [mh ^"greenhouse effect"] | 2 |
| #11 | [mh ^"extreme heat"] | 6 |
| #12 | [mh ^"hot temperature"] | 1868 |
| #13 | [mh ^"carbon footprint"] | 2 |
| #14 | (climat* near/3 (chang* or disruption* or instabilit* or stabilit* or stable or unstable or variable or variabilit* or vulnerab*)):ti,ab,kw | 140 |
| #15 | (weather near/3 (severe or chang* or disruption* or instabilit* or stabilit* or stable or unstable or variable or variabilit* or vulnerab*)):ti,ab,kw | 42 |
| #16 | (environment* near/3 (chang* or disruption* or instabilit* or stabilit* or stable or unstable or variable or variabilit* or vulnerab*)):ti,ab,kw | 891 |

| | | |
|---|---|---|
| #17 | ((chang* or declin* or decreas* or increas*) near/3 humid*) | 134 |
| #18 | ((global* or climate) near/2 (warm* or heat*)):ti,ab,kw | 60 |
| #19 | "climatic condition*":ti,ab,kw | 4 |
| #20 | "carbon footprint*":ti,ab,kw | 17 |
| #21 | ("greenhouse gas*" or GHG or "greenhouse effect*" or "carbon emission*" or "carbon dioxide emission*" or "CO2 emission*"):ti,ab,kw | 55 |
| #22 | ("extreme weather" or "extreme heat" or "hot weather" or "heat wave*" or heatwave or ((high* or hot* or warm*) near/3 temperature*)):ti,,ab,kw | 3416 |
| #23 | (rain or rains or raining or precipitation or "UV index" or flooding or floods or waterlogging or drought* or desertifcation or storm* or hailstorm* or hail or hailing* or monsoon* or typhoon*):ti,ab,kw | 1385 |
| #24 | ("ice storm*" or blizzard* or snowstorm* or "extreme cold*" or "polar vortex*"):ti,ab,kw | 18 |
| #25 | (((high* or extreme) adj2 wind*) or hurricane* or cyclone* or tornado* or waterspout*):ti,ab,kw | 92 |
| #26 | ("forest fire*" or wildfire* or "wild fire*"):ti,ab,kw | 14 |
| #27 | ("sea level*" near/3 (rise or rises or rising or increase or increasing or increases)):ti,ab,kw | 8 |
| #28 | {or #1-#27} with Cochrane Library publication date Between Jan 1999 and Jul 2020 | 5943 (200 reviews & protocols – kept) |

| |
|---|
| **Database: ProQuest Dissertations & Theses Global** |
| *NOTE: ProQuest could not handle adding the new additional review terminology to the search; had to break into separate searches and then combine/deduplicate.* |

su("climate change" OR "weather" OR "climate" OR "Temperature" OR "climatic process*" OR "cyclonic storm*" OR "hurricane*" OR "drought*" OR "flood*" OR "greenhouse effect" OR "extreme heat" OR "hot temperature*" OR "carbon footprint*") OR noft((climat* near/3 (chang* or disruption* or instabilit* or stabilit* or stable or unstable or variable or variabilit* or vulnerab*)) or (weather near/3 (chang* or disruption* or instabilit* or stabilit* or stable or unstable or variable or variabilit* or vulnerab*)) or (environment* near/3 (chang* or disruption* or instabilit* or stabilit* or stable or unstable or variable or variabilit* or vulnerab*)) or ((chang* or declin* or decreas* or increas*) near/3 humid*) or ((global* or climate) near/2 (warm* or heat*)) or "climatic condition*" or "carbon footprint*" or "greenhouse gas*" or GHG or "greenhouse effect*" or "carbon emission*" or "carbon dioxide emission*" or "CO2 emission*" or "extreme weather" or "extreme heat" or "hot weather" or "heat wave*" or heatwave or ((high* or hot* or warm*) near/3 temperature*) or rain or rains or raining or precipitation or "UV index" or flooding or floods or waterlogging or drought* or desertifcation or storm* or hailstorm* or hail or hailing* or monsoon* or typhoon* or "ice storm*" or blizzard* or snowstorm* or "extreme cold*" or "polar vortex*" or ((high* or extreme) near/2 wind*) or hurricane* or cyclone* or tornado* or waterspout* or "forest fire*" or wildfire* or "wild fire*" or ("sea level*" near/3 (rise or rises or rising or increase or increasing or increases)) )

AND

(su((literature near/3 review*) or scoping or ("technical briefings") ) OR noft((scoping near/3 (review* or overview* or search* or study or studies or exercise* or project* or report* or methodolog*)) or ((scope or scoping) near/2 evidence) or ("technical briefings") or ((evidence or systematic or literature or review* or overview*) near/3 map*) or (structured near/2 ("literature review" OR "literature reviews")) or ((research or literature) near/3 (gap or gaps))))

OR

(su(meta-analys* OR metaanalys* OR "technolog* assess*") OR noft((quantitativ* NEAR/3 review*) OR (quantitativ* NEAR/3 overview*) OR (quantitativ* NEAR/3 synthes*) OR "data synthes*" OR "data extraction*" OR "data abstraction*" OR (systematic* NEAR/3 review*) OR (systematic* NEAR/3 overview*) OR (methodologic NEAR/3 review*))) OR noft((methodologic NEAR/3 overview*) OR (integrat* NEAR/5 research) OR (integrative NEAR/3 review*) OR (integrative NEAR/3 overview*) OR (research NEAR/3 integrati*) OR (research NEAR/3 overview) OR (collaborative NEAR/3 review*) OR (collaborative NEAR/3 overview*) OR (overview NEAR/2 review*) OR (rapid* NEAR/2 review*) OR (Technology NEAR/1 assessment*) OR (technology NEAR/1 overview*) OR (technology NEAR/1 appraisal*))

Narrowed by:
Entered date: 1999-01-01 - 2021-02-28; Language: English; Total: 1676; 1623 w/duplicates removed

### Appendix 2: Included evidence syntheses sorted by number of included studies

| Extracted | Included studies | Database ID | Year | Title | Evidence synthesis | Journal | Pathways |
|---|---|---|---|---|---|---|---|
| **study extracted LSW** | 6 | 2004172075 | 2020 | A systematic review of respiratory infection due to air pollution during natural disasters | Sys Rev or Scop Rev | Medical Journal of Indonesia | air quality |
| **study extracted MIM** | 7 | 31884209 | 2020 | Environmental temperature and human epigenetic modifications: A systematic review | Sys Rev or Scop Rev | Environmental Pollution | heat stress |
| **study extracted MIM** | 8 | 30675732 | 2019 | Evaluation of the impact of heat stress on the occurrence of occupational injuries: Meta-analysis of observational studies | Meta-Analysis | American Journal of Industrial Medicine | heat stress |
| **study extracted LSW** | 9 | 25557350 | 2015 | Climate Variability and the Occurrence of Human Puumala Hantavirus Infections in Europe: A Systematic Review | Sys Rev or Scop Rev | Zoonoses & Public Health | vector distribution and ecology |
| **study extracted DT** | 9 | 26931438 | 2016 | Research on Climate and Dengue in Malaysia: A Systematic Review | Sys Rev or Scop Rev | Current Environmental Health Reports | vector distribution and ecology |
| **study extracted DT** | 10 | 30677927 | 2019 | What do we know about the healthcare costs of extreme heat exposure? A comprehensive literature review | Comprehensive Lit Rev | Science of the Total Environment | heat stress |
| **study extracted IM** | 10 | 30889810 | 2019 | Meta-Analysis of Heterogeneity in the Effects of Wildfire Smoke Exposure on Respiratory Health in North America | Meta-Analysis | International Journal of Environmental Research & Public Health | air quality |
| **study extracted KJ** | 11 | 32843012 | 2020 | Health effects of heating, ventilation and air conditioning on hospital patients: a scoping review | Sys Rev or Scop Rev | BMC Public Health | heat stress |
| **study extracted DT** | 12 | 23525899 | 2014 | The impact of heat waves on children's health: a systematic review | Sys Rev or Scop Rev | International Journal of Biometeorology | heat stress |
| **study extracted LSW** | 12 | 25563349 | 2015 | Climate change, water quality, and water-related diseases in the Mekong Delta Basin: a systematic review | Sys Rev or Scop Rev | Asia-Pacific Journal of Public Health | water quality and quantity |

| | | | | | | | |
|---|---|---|---|---|---|---|---|
| **study extracted DT** | 13 | 26868947 | 2016 | Impact of high ambient temperature on unintentional injuries in high-income countries: a narrative systematic literature review | Sys Rev or Scop Rev | BMJ Open | heat stress |
| **study extracted IM** | 13 | 619966140 | 2017 | The effect of climate change on cardiopulmonary disease-a systematic review | Sys Rev or Scop Rev | Journal of Clinical and Diagnostic Research | air quality |
| **study extracted MIM** | 14 | 21816703 | 2011 | Projecting future heat-related mortality under climate change scenarios: a systematic review | Sys Rev or Scop Rev | Environmental Health Perspectives | heat stress |
| **study extracted IM** | 14 | 26908518 | 2016 | Long-term exposure to ambient ozone and mortality: a quantitative systematic review and meta-analysis of evidence from cohort studies | Sys Rev or Scop Rev | BMJ Open | air quality |
| **study extracted IM** | 14 | 29331087 | 2018 | Outdoor pollen is a trigger of child and adolescent asthma emergency department presentations: A systematic review and meta-analysis | Sys Rev or Scop Rev | Allergy | air quality |
| **study extracted IM** | 14 | 29428708 | 2018 | Surface water flooding, groundwater contamination, and enteric disease in developed countries: A scoping review of connections and consequences | Sys Rev or Scop Rev | Environmental Pollution | water quality and quantity |
| **study extracted LSW** | 14 | 30166779 | 2018 | A systematic evidence review of the effect of climate change on malaria in Iran | Sys Rev or Scop Rev | Journal of Parasitic Diseases | vector distribution and ecology |
| **study extracted IM** | 15 | 19079707 | 2008 | Climate variability, social and environmental factors, and ross river virus transmission: research development and future research needs | Sys Search' (in abstract) | Environmental Health Perspectives | vector distribution and ecology |
| **study extracted MIM** | 15 | 21975970 | 2012 | Daily average temperature and mortality among the elderly: a meta-analysis and systematic review of epidemiological evidence | Sys Rev or Scop Rev | International Journal of Biometeorology | heat stress |
| **study extracted MIM** | 15 | 25461412 | 2015 | Projecting future air pollution-related mortality under a changing climate: progress, uncertainties and research needs | Sys Rev or Scop Rev (in abstract) | Environment International | air quality |
| **study extracted LSW** | 15 | 26109998 | 2015 | Systematic review on adverse birth outcomes of climate change | Sys Rev or Scop Rev | Journal of Research in Medical Sciences | heat stress |

| | | | | | | | |
|---|---|---|---|---|---|---|---|
| **study extracted DT** | 15 | 26216952 | 2015 | Systematic review of current efforts to quantify the impacts of climate change on undernutrition | Sys Rev or Scop Rev | Proceedings of the National Academy of Sciences of the United States of America | food supply and safety |
| **study extracted IM** | 15 | 27918457 | 2016 | Air Quality Strategies on Public Health and Health Equity in Europe-A Systematic Review | Sys Rev or Scop Rev | International Journal of Environmental Research & Public Health | air quality |
| **study extracted DT** | 15 | 32228631 | 2020 | Heat-health vulnerability in temperate climates: lessons and response options from Ireland | Sys Rev or Scop Rev (in abstract) | Global Health | heat stress |
| **study extracted IM** | 16 | 24669859 | 2014 | Climate change and dengue: a critical and systematic review of quantitative modelling approaches | Sys Rev or Scop Rev | BMC Infectious Diseases | vector distribution and ecology |
| **study extracted KJ** | 16 | 30177247 | 2018 | Climate change and sleep: A systematic review of the literature and conceptual framework | Sys Rev or Scop Rev | Sleep Medicine Reviews | multiple pathways: extreme weather events; heat stress; air quality; social factors |
| **study extracted IM** | 16 | 32119666 | 2020 | Projecting the future of dengue under climate change scenarios: Progress, uncertainties and research needs | Sys Rev or Scop Rev (in abstract) | PLoS Neglected Tropical Diseases | vector distribution and ecology |
| **study extracted MIM** | 16 | 33168031 | 2020 | Climate factors and gestational diabetes mellitus risk - a systematic review | Sys Rev or Scop Rev | Environmental Health | multiple pathways: heat stress; extreme weather events |
| **study extracted IM** | 17 | 27682833 | 2016 | Extreme weather events in developing countries and related injuries and mental health disorders - a systematic review | Sys Rev or Scop Rev | BMC Public Health | extreme weather events |
| **study extracted IM** | 17 | 31169886 | 2020 | Natural disasters and infectious disease in Europe: a literature review to identify cascading risk pathways | Sys Rev or Scop Rev (in abstract) | European Journal of Public Health | multiple pathways: extreme weather events; vector distribution and ecology; food suply and safety; water quality and quantity |
| **study extrected IM** | 17 | 32876770 | 2020 | Climate and climate-sensitive diseases in semi-arid regions: a systematic review | Sys Rev or Scop Rev | International Journal of Public Health | multiple pathways: extreme weather events; vector distribution and ecology; water quality and quantity; air quality |
| **study extracted IM** | 17 | 33227944 | 2020 | Effect of Extreme Weather Events on Mental Health: A Narrative Synthesis and Meta-Analysis for the UK | Meta-Analysis | International Journal of Environmental Research & Public Health | extreme weather events |

| | | | | | | | |
|---|---|---|---|---|---|---|---|
| **study extracted IM** | 17 | 130177110 | 2018 | Public Health Impacts of heat waves: A Review | Sys Search' (in abstract) | International Journal of Public Health & Clinical Sciences (IJPHCS) | heat stress |
| **study extracted LSW** | 18 | 31717424 | 2019 | Heat Health Prevention Measures and Adaptation in Older Populations-A Systematic Review | Sys Rev or Scop Rev | International Journal of Environmental Research & Public Health | heat stress |
| **study extracted IM** | 19 | 29022096 | 2018 | The association between ambient temperature and childhood asthma: a systematic review | Sys Rev or Scop Rev | International Journal of Biometeorology | heat stress |
| **study extracted RK** | 19 | 33415391 | 2021 | Lymphatic filariasis in Asia: a systematic review and meta-analysis | Sys Rev or Scop Rev | Parasitology Research | vector distribution and ecology |
| **study extracted MIM** | 20 | 24459613 | 2014 | Power outages, extreme events and health: a systematic review of the literature from 2011-2012 | Sys Rev or Scop Rev | PLoS currents | extreme weather events |
| **study extracted IM** | 20 | 27834843 | 2016 | Economic Evaluations of the Health Impacts of Weather-Related Extreme Events: A Scoping Review | Sys Rev or Scop Rev | International Journal of Environmental Research & Public Health | extreme weather events |
| **study extracted IM** | 20 | 32509905 | 2020 | Migration health crisis associated with climate change: A systematic review | Sys Rev or Scop Rev | Journal of Education & Health Promotion | social factors |
| **study extracted IM** | 22 | 26309294 | 2014 | Ross River Virus Disease activity associated with naturally occurring nontidal flood events in Australia: A Systematic Review | Sys Rev or Scop Rev | Journal of Medical Entomology | vector distribution and ecology |
| **study extracted IM** | 22 | 33332222 | 2020 | A systematic review and meta-analysis assessing the impact of droughts, flooding, and climate variability on malnutrition | Sys Rev or Scop Rev | Global Public Health | multiple pathways: food supply and safety; extreme weather events |
| **study extracted IM** | 23 | 26949867 | 2016 | Temperature-related morbidity and mortality in Sub-Saharan Africa: A systematic review of the empirical evidence | Sys Rev or Scop Rev | Environment International | multiple pathways: heat stress; air quality , water quality and quantity, food supply and safety |

| | | | | | | | |
|---|---|---|---|---|---|---|---|
| **study extracted IM** | 24 | 33472088 | 2021 | Extreme heat and occupational injuries in different climate zones: A systematic review and meta-analysis of epidemiological evidence | Sys Rev or Scop Rev | Environment International | heat stress |
| **study extracted IM** | 24 | 2004325232 | 2020 | Investigating the resurgence of malaria prevalence in South Africa between 2015 and 2018: A scoping review | Sys Rev or Scop Rev | Open Public Health Journal | vector distribution and ecology |
| **study extracted IM** | 26 | 26567313 | 2016 | A systematic review and meta-analysis of ambient temperature and diarrhoeal diseases | Sys Rev or Scop Rev | International Journal of Epidemiology | multiple pathways: water quality and quantity; food supply and safety; vector distribution and ecology; extreme weather events |
| **study extracted IM** | 26 | 32823825 | 2020 | Drought Influences on Food Insecurity in Africa: A Systematic Literature Review | Sys Rev or Scop Rev | International Journal of Environmental Research & Public Health | multiple pathways: extreme weather events; food supply and safety |
| **study extracted KJ** | 26 | 32965555 | 2021 | Global climate implications for homelessness: A scoping review | Sys Rev or Scop Rev | Journal of Urban Health | social factors |
| **study extracted IM** | 26 | 2000775241 | 2018 | Are workers at risk of occupational injuries due to heat exposure? A comprehensive literature review | Comprehensive Lit Rev | Safety Science | heat stress |
| **not extracted** | 26 | 28717366 | 2017 | Interrelationship between Climatic, Ecologic, Social, and Cultural Determinants Affecting Dengue Emergence and Transmission in Puerto Rico and Their Implications for Zika Response | Sys Rev or Scop Rev (in abstract) | Journal of Tropical Medicine | vector distribution and ecology |
| **study extracted IM** | 27 | 31400736 | 2019 | Drought exposure as a risk factor for child undernutrition in low- and middle-income countries: A systematic review and assessment of empirical evidence | Sys Rev or Scop Rev | Environment International | food supply and safety |
| **study extracted IM** | 27 | 31425512 | 2019 | Distribution of geographical scale, data aggregation unit and period in the correlation analysis between temperature and incidence of HFRS in mainland China: A systematic review of 27 ecological studies | Sys Rev or Scop Rev | PLoS Neglected Tropical Diseases | vector distribution and ecology |

| | | | | | | | |
|---|---|---|---|---|---|---|---|
| **study extracted IM** | 27 | 33395923 | 2021 | Association between ambient temperature and heat waves with mortality in South Asia: Systematic review and meta-analysis | Sys Rev or Scop Rev | Environment International | heat stress |
| **study extracted DT** | 28 | 26931446 | 2016 | Climatic Drivers of Diarrheagenic Escherichia coli Incidence: A Systematic Review and Meta-analysis | Sys Rev or Scop Rev | Journal of Infectious Diseases | water quality and quantity |
| **not extracted** | 28 | 28758917 | 2017 | Heat Exposure and Maternal Health in the Face of Climate Change | Sys Rev or Scop Rev (in abstract) | International Journal of Environmental Research & Public Health | heat stress |
| **study extracted IM** | 29 | 24990685 | 2014 | Association of climatic factors with infectious diseases in the Arctic and subarctic region--a systematic review | Sys Rev or Scop Rev | Glob Health Action | multiple pathways: water quality and quantity; food supply and safety; vector distribution and ecology |
| **study extracted KJ** | 29 | 31349659 | 2019 | Mapping the Solastalgia Literature: A Scoping Review Study | Sys Rev or Scop Rev | International Journal of Environmental Research & Public Health | multiple pathways: social factors |
| **study extracted IM** | 29 | 32584659 | 2020 | The impact of climate change on mosquito-borne diseases in Africa | Sys Rev or Scop Rev (in abstract) | Pathogens and Global Health | vector distribution and ecology |
| **study extracted KJ** | 29 | 33284688 | 2020 | Management Of Chronic Noncommunicable Diseases After Natural Disasters In The Caribbean: A Scoping Review | Sys Rev or Scop Rev | Health Affairs | extreme weather events |
| **not extracted** | 30 | 25349109 | 2014 | Is planned adaptation to heat reducing heat-related mortality and illness? A systematic review | Sys Rev or Scop Rev | BMC Public Health | heat stress |
| **study extracted IM** | 30 | 30029319 | 2018 | Effects of ambient temperature on myocardial infarction: A systematic review and meta-analysis | Sys Rev or Scop Rev | Environmental Pollution | heat stress |
| **study extracted HJ** | 30 | 32119682 | 2020 | Seroprevalence of Crimean-Congo hemorrhagic fever in humans in the World Health Organization European region: A systematic review | Sys Rev or Scop Rev | PLoS Neglected Tropical Diseases | vector distribution and ecology |
| **not extracted** | 33 | 22831555 | 2012 | Impact of ambient temperature on children's health: a systematic review | Sys Rev or Scop Rev | Environmental Research | heat stress |

| | | | | | | | |
|---|---|---|---|---|---|---|---|
| **not extracted** | 33 | 26900154 | 2015 | The Effects of Climate Change on Patients With Chronic Lung Disease. A Systematic Literature Review | Sys Rev or Scop Rev | Deutsches Arzteblatt International | heat stress |
| **study extracted IM** | 34 | 30200277 | 2018 | Impacts of Climate Change on Health and Wellbeing in South Africa | We systematically reviewed the literature' | International Journal of Environmental Research & Public Health | multiple pathways: extreme weather events; heat stress; water quality and quantity; food supply and safety; vector distribution and ecology; social factors |
| **study extracted IM** | 35 | 33530011 | 2021 | Extreme weather events in europe and their health consequences - A systematic review | Sys Rev or Scop Rev | International Journal of Hygiene & Environmental Healt | extreme weather events |
| **not extracted** | 36 | 33515577 | 2021 | The effect of the heatwave on the morbidity and mortality of diabetes patients; a meta-analysis for the era of the climate crisis | Meta-Analysis | Environmental Research | heat stress |
| **not extracted** | 37 | 25493706 | 2015 | The potential impact of climate change and ultraviolet radiation on vaccine-preventable infectious diseases and immunization service delivery system | We systematically reviewed the scientific literature' | Expert Review of Vaccines | vector distribution and ecology |
| **study extracted KJ** | 37 | 25503413 | 2014 | Impact of ambient humidity on child health: a systematic review | Sys Rev or Scop Rev | PLoS ONE | air quality |
| **study extracted IM** | 38 | 28228994 | 2013 | Examining the relationship between infectious diseases and flooding in Europe: A systematic literature review and summary of possible public health interventions | Sys Rev or Scop Rev | Disaster Health | multiple pathways: extreme weather; water quality and quantity; vector distributuion and ecology |
| **not extracted** | 40 | 31340512 | 2019 | Scoping Review of Climate Change and Health Research in the Philippines: A Complementary Tool in Research Agenda-Setting | Sys Rev or Scop Rev | International Journal of Environmental Research & Public Health | multiple pathways: vector distribution and ecology; air quality; extreme weather events; water quality and quantity; food supply and safety; heat stress |
| **not extracted** | 43 | 30317100 | 2018 | Human infectious diseases and the changing climate in the Arctic | Sys Rev or Scop Rev (in abstract) | Environment International | vector distribution and ecology |

| | | | | | | | |
|---|---|---|---|---|---|---|---|
| **study extracted IM** | 43 | 32661979 | 2020 | How climate change can affect cholera incidence and prevalence? A systematic review | Sys Rev or Scop Rev | Environmental Science & Pollution Research | water quality and quantity |
| **study extracted DT** | 47 | 28834176 | 2017 | Climate Change-Related Water Disasters' Impact on Population Health | Sys Rev or Scop Rev (in abstract) | Journal of Nursing Scholarship | extreme weather events |
| **study extracted IM** | 50 | 26086887 | 2015 | Climate Change and Spatiotemporal Distributions of Vector-Borne Diseases in Nepal--A Systematic Synthesis of Literature | Sys Search (in abstract) | PLoS ONE | vector distribution and ecology |
| **not extracted** | 51 | 30451130 | 2018 | Meteorological factors and its association with hand, foot and mouth disease in Southeast and East Asia areas: a meta-analysis | Meta-Analysis | Epidemiology & Infection | vector distribution and ecology |
| **study extracted IM** | 53 | 32300088 | 2020 | Water, sanitation and hygiene risk factors for the transmission of cholera in a changing climate: using a systematic review to develop a causal process diagram | Sys Rev or Scop Rev | Journal of Water & Health | water quality and quantity |
| **study extracted CEL** | 53 | 612021889 | 2016 | Critical review of health impacts of wildfire smoke exposure | Critical Review | Environmental Health Perspectives | air quality |
| **study extracted IM** | 56 | 33287833 | 2020 | Synergistic health effects of air pollution, temperature, and pollen exposure: a systematic review of epidemiological evidence | Sys Rev or Scop Rev | Environmental Health: A Global Access Science Source | multiple pathways: heat stress; air quality |
| **not extracted** | 60 | 26878285 | 2016 | Impact of heatwave on mortality under different heatwave definitions: A systematic review and meta-analysis | Sys Rev or Scop Rev | Environment International | heat stress |
| **not extracted** | 60 | 27211569 | 2016 | Effects of Air Temperature on Climate-Sensitive Mortality and Morbidity Outcomes in the Elderly; a Systematic Review and Meta-analysis of Epidemiological Evidence | Sys Rev or Scop Rev | EBioMedicine | heat stress |
| **study extracted LSW** | 61 | 25460628 | 2015 | A systematic review of the physical health impacts from non-occupational exposure to wildfire smoke | Sys Rev or Scop Rev | Environmental Research | air quality |

| | | | | | | | |
|---|---|---|---|---|---|---|---|
| **study extracted IM** | 61 | 32810503 | 2020 | Environmental drivers, climate change and emergent diseases transmitted by mosquitoes and their vectors in southern Europe: A systematic review | Sys Rev or Scop Rev | Environmental Research | vector distribution and ecology |
| **not extracted** | 63 | 28686743 | 2017 | The use of climate information to estimate future mortality from high ambient temperature: A systematic literature review | Sys Rev or Scop Rev | PLoS ONE | heat stress |
| **study extracted IM** | 65 | 24907712 | 2014 | The effects of season and meteorology on human mortality in tropical climates: a systematic review | Sys Rev or Scop Rev | Transactions of the Royal Society of Tropical Medicine & Hygiene | multiple pathways: extreme weather events; heat stress; water quality and quantity |
| **study extracted CEL** | 68 | 32556259 | 2020 | Association of Air Pollution and Heat Exposure With Preterm Birth, Low Birth Weight, and Stillbirth in the US: A Systematic Review | Sys Rev or Scop Rev | JAMA Network Open | multiple pathways: air quality; heat stress |
| **study extracted IM, KJ, DT** | 68 | 33327439 | 2020 | A Meta-Synthesis of Policy Recommendations Regarding Human Mobility in the Context of Climate Change | Sys Search' (in abstract) | International Journal of Environmental Research & Public Health | social factors |
| **not extracted** | 69 | 32102956 | 2019 | A Scoping Review of Nurses' Contributions to Health-Related, Wildfire Research | Sys Rev or Scop Rev | Annual Review of Nursing Research | extreme weather events; air quality |
| **not extracted** | 70 | 33148618 | 2020 | Associations between high temperatures in pregnancy and risk of preterm birth, low birth weight, and stillbirths: systematic review and meta-analysis | Sys Rev or Scop Rev | BMJ | heat stress |
| **not extracted** | 70 | 33508094 | 2021 | The impact of climate change on neglected tropical diseases: a systematic review | Sys Rev or Scop Rev | Transactions of the Royal Society of Tropical Medicine & Hygiene | vector distribution and ecology |
| **not extracted** | 72 | 30187452 | 2019 | Temperature and humidity affect the incidence of hand, foot, and mouth disease: a systematic review of the literature - a report from the International Society of Dermatology Climate Change Committee | Sys Rev or Scop Rev | International Journal of Dermatology | vector distribution and ecology |

| | | | | | | | |
|---|---|---|---|---|---|---|---|
| **study extracted IM** | 78 | 26351799 | 2015 | Climate change and health in the Eastern Mediterranean countries: a systematic review | Sys Rev or Scop Rev | Reviews on Environmental Health | multiple pathways: extreme weather events; air quality; vector distrubution and ecology; social factors; food supply and safety; water quality and quantity; heat stress |
| **not extracted** | 81 | 29220773 | 2018 | Climate change and dengue fever transmission in China: Evidences and challenges | Sys Rev or Scop Rev | Science of the Total Environment | vector distribution and ecology |
| **not extracted** | 81 | 29907109 | 2018 | Epidemiological trends and risk factors associated with dengue disease in Pakistan (1980-2014): a systematic literature search and analysis | Sys Search' (in abstract) | BMC Public Health | vector distribution and ecology |
| **study extracted IM** | 83 | 22877498 | 2013 | Extreme water-related weather events and waterborne disease | Sys Rev or Scop Rev (in abstract) | Epidemiology & Infection | extreme weather events |
| **study extracted IM** | 87 | 23787891 | 2013 | Health effects of drought: a systematic review of the evidence | Sys Rev or Scop Rev | PLoS currents | extreme weather events |
| **study extracted HJ** | 91 | 30942769 | 2019 | Salmonella and the changing environment: systematic review using New York State as a model | Sys Rev or Scop Rev | Journal of Water & Health | food supply and safety |
| **not extracted** | 97 | 31011886 | 2019 | Impacts of exposure to ambient temperature on burden of disease: a systematic review of epidemiological evidence | Sys Rev or Scop Rev | International Journal of Biometeorology | heat stress |
| **not extracted** | 111 | 30526938 | 2018 | Workers' health and productivity under occupational heat strain: a systematic review and meta-analysis | Sys Rev or Scop Rev | The Lancet. Planetary Health | heat stress |
| **not extracted** | 112 | 33119678 | 2020 | The effect of air-pollution and weather exposure on mortality and hospital admission and implications for further research: A systematic scoping review | Sys Rev or Scop Rev | PLoS ONE | multiple pathways: air quality; heat stress; extreme weather events |
| **study extracted IM** | 141 | 27058059 | 2016 | Untangling the Impacts of Climate Change on Waterborne Diseases: a Systematic Review of Relationships between Diarrheal Diseases and Temperature, Rainfall, Flooding, and Drought | Sys Rev or Scop Rev | Environmental Science & Technology | water quality and quantity |

| | | | | | | | |
|---|---|---|---|---|---|---|---|
| **not extracted** | 162 | 32914743 | 2020 | Drivers and health implications of the dietary transition among Inuit in the Canadian Arctic: a scoping review | Sys Rev or Scop Rev | Public Health Nutrition | food supply and safety |
| **study extracted HJ (only "extreme weather events")** | 163 | 32210846 | 2020 | The Impact of Climate Change on Mental Health: A Systematic Descriptive Review | Sys Rev or Scop Rev | Frontiers in psychiatry Frontiers Research Foundation | multiple pathways: extreme weather events; social factors; heat stress; vector distribution and ecology |
| **not extracted** | 175 | 32833174 | 2020 | A review of the impact of outdoor and indoor environmental factors on human health in China | Sys Rev or Scop Rev (in abstract) | Environmental Science & Pollution Research | multiple pathways: air quality; heat stress; extreme weather events; vector distribution and ecology |
| **not extracted** | 178 | 29911344 | 2018 | Powassan virus, a scoping review of the global evidence | Sys Rev or Scop Rev | Zoonoses & Public Health | vector distribution and ecology |
| **not extracted** | 193 | 29943748 | 2018 | Impact of climate change on occupational health and productivity | Sys Rev or Scop Rev (in abstract) | Medicina del Lavoro | heat stress |
| **not extracted** | 320 | 18400023 | 2008 | Does climate change affect the incidence of skin and allergic diseases in Germany? | Sys Search' (in abstract) | Journal der Deutschen Dermatologischen Gesellschaft | air quality |
| **not extracted** | 380 | 33129458 | 2020 | Marine harmful algal blooms and human health: A systematic scoping review | Sys Rev or Scop Rev | Harmful Algae | water quality and quantity |
| **not extracted** | 428 | 31940405 | 2020 | A scoping review of importation and predictive models related to vector-borne diseases, pathogens, reservoirs, or vectors (1999-2016) | Sys Rev or Scop Rev | BMJ Open | vector distribution and ecology |
| **not extracted** | 1394 | 31887120 | 2019 | Zoonotic Babesia: A scoping review of the global evidence | Sys Rev or Scop Rev | PLoS ONE | vector distribution and ecology |
| **not extracted** | 1920 | 30496207 | 2018 | A scoping review of published literature on chikungunya virus | Sys Rev or Scop Rev | PLoS ONE | vector distribution and ecology |

**Appendix 3: Search filter translation to Ovid MEDLINE syntax**

| | 95% sensitivity cutoff | 97% sensitivity cutoff | 99% sensitivity cutoff |
|---|---|---|---|
| **Air quality** | | | |
| **Filter** | exp Fires/ or exp Particulate Matter/ or Smoke/ or (wildfire* or PM10 or "PM 10" or PM2 or air pollution or air quality or "PM 2.5" or ozone or weather or pollutant* or humid*).ti,ab. | exp Fires/ or exp Particulate Matter/ or exp Air Pollution/ or Smoke/ or (wildfire* or PM10 or "PM 10" or PM2 or air pollution or air quality or "PM 2.5" or ozone or weather or pollutant* or humid* or ambient).ti,ab. | exp Fires/ or exp Particulate Matter/ or exp Air Pollution/ or exp Smoke/ or (wildfire* or PM10 or PM 10 or PM2 or air pollution or air quality or "PM 2.5" or ozone or weather or pollutant* or humid* or ambient or asthma).ti,ab. |
| **Extreme weather events** | | | |
| **Filter** | Floods/ or Cyclonic Storms/ or exp Disasters/ or (river* or hurricane* or flood* or disaster* or drought* or weather).ti,ab. | Floods/ or Cyclonic Storms/ or exp Disasters/ or (river* or hurricane* or flood* or disaster* or drought* or weather or rainfall*).ti,ab. | Floods/ or Cyclonic Storms/ or exp Disasters/ or exp Climate/ or (river* or hurricane* or flood* or disaster* or drought* or weather or rainfall*).ti,ab. |
| **Food supply and safety** | | | |
| **Filter** | Child Nutrition Disorders/ or Cholera/ or exp Rain/ or exp Food Supply/ or Droughts/ or exp Salmonella Infections/ or exp Foodborne Diseases/ or (salmonellos* or stunt* or drought* or underweight or foodborne or malnutrition or weather).ti,ab. | Child Nutrition Disorders/ or Cholera/ or exp Rain/ or exp Food Supply/ or Droughts/ or exp Salmonella Infections/ or exp Foodborne Diseases/ or (salmonellos* or stunt* or drought* or underweight or foodborne or malnutrition or weather or salmonella*).ti,ab. | Child Nutrition Disorders/ or Cholera/ or exp Rain/ or exp Food Supply/ or Droughts/ or exp Salmonella Infections/ or exp Foodborne Diseases/ or Food Microbiology/ or Seasons/ or (salmonellos* or stunt* or drought* or underweight or foodborne or malnutrition or weather or salmonella*).ti,ab. |
| **Heat stress** | | | |

| | | | |
|---|---|---|---|
| **Filter** | Extreme Heat/ or exp Heat Stress Disorders/ or exp Hot Temperature/ or Seasons/ or (daily mortality or heat wave* or heat related or extreme heat or heatwave* or weather or meteorologic* or ambient temperature* or climate change or air pollution or summer*).ti,ab. | Extreme Heat/ or exp Heat Stress Disorders/ or exp Hot Temperature/ or Seasons/ or (daily mortality or heat wave* or heat related or extreme heat or heatwave* or weather or meteorologic* or ambient temperature* or climate change or air pollution or summer* or climate* or hot).ti,ab. | Extreme Heat/ or exp Heat Stress Disorders/ or exp Hot Temperature/ or Seasons/ or exp Climate/ or (daily mortality or heat wave* or heat related or extreme heat or heatwave* or weather or meteorologic* or ambient temperature* or climate change or air pollution or summer* or climate* or hot or high temperature* or ambient).ti,ab. |
| **Vector distribution and ecology** | | | |
| **Filter** | Rain/ or Climate/ or Insect Vectors/ or Climate Change/ or Seasons/ or transmission.fs. or Disease Outbreaks/ or (rain* or mosquito* or relative humidity or climat* or borne or endemic* or fever).ti,ab. | Rain/ or Climate/ or Insect Vectors/ or Climate Change/ or Seasons/ or transmission.fs. or Disease Outbreaks/ or (rain* or mosquito* or relative humidity or climat* or borne or endemic* or epidemic* or fever).ti,ab. | Rain/ or Climate/ or Insect Vectors/ or Climate Change/ or Seasons/ or transmission.fs. or Disease Outbreaks/ or (rain* or mosquito* or climat* or humi* or borne or endemic* or epidemic* or fever or vector* or surveillan*).ti,ab. |
| **Water quality and quantity** | | | |
| **Filter** | Cholera/ or Floods/ or exp Vibrio cholerae/ or Water Microbiology/ or exp Water Supply/ or exp Climate/ or (heavy rain* or waterborne or rainfall* or flood* or diarrheal or cholera* or diarrhoea*).ti,ab. | Cholera/ or Floods/ or exp Vibrio cholerae/ or Water Microbiology/ or exp Water Supply/ or exp Diarrhea/ or exp Climate/ or (heavy rain* or waterborne or rainfall* or flood* or diarrh* or cholera*).ti,ab. | Cholera/ or Floods/ or exp Vibrio cholerae/ or Water Microbiology/ or exp Water Supply/ or exp Disasters/ or exp Climate/ or (heavy rain* or waterborne or rainfall* or flood* or diarrh* or cholera* or season*).ti,ab. |
